# Consumer continuous glucose monitoring uncovers hidden metabolic risk in free-living adults without diabetes

**DOI:** 10.64898/2026.11.03.26367126

**Authors:** Paul Ginart, Michael Loesche

## Abstract

Whether continuous glucose monitoring (CGM) provides actionable information in adults without diabetes is contested, as its metrics were designed for diabetes and screening relies on HbA1c. In this cross-sectional study of 62,122 free-living adults on the Levels digital-health platform, average glucose was the most reliable measure across weeks (intraclass correlation 0.747). Among 7,930 participants without diabetes with laboratory data, it tracked insulin resistance, triglycerides, HDL cholesterol, uric acid, and C-reactive protein more closely than HbA1c, explaining nearly three times more variance in fasting insulin (R² 0.131 vs 0.048). With a normal HbA1c, occult hyperinsulinemia (fasting insulin ≥10 µIU/mL) was twice as common at average glucose ≥110 mg/dL (27.5% vs 12.6%). Counterintuitively, at the same average glucose, spike width, not height or frequency, tracked insulin resistance, triglycerides, and C-reactive protein; carbohydrate sensitivity, although reliable, did not track insulin resistance. This analysis suggests a framework for interpreting CGM in adults without diabetes.

## Introduction

Human health is becoming continuously observable. Wearable sensors, app-logged behavior, and direct-to-consumer laboratory testing now capture human physiology in daily life over months to years, rather than at isolated clinic visits.^1,2^ Continuous glucose monitors (CGMs) are at the forefront of this shift. Cleared for over-the-counter sale in the United States in 2024,^3^ CGMs are increasingly worn by adults without diabetes to track their metabolic health and personalize their diets,^4^ while AI foundation models are being trained on the resulting traces to infer cardiometabolic risk.^5,6^

Yet there is no clinical consensus on whether, or how, to use CGM in people without diabetes.^7,8^ Because conventional CGM targets such as time in range and the coefficient of variation (CV) were derived in populations with diabetes,^9^ many clinicians question whether CGM adds actionable information beyond a standard HbA1c,^7,8,10^ and experts disagree on how to interpret reports in this population.^11^ Resolving this debate requires determining which features of a non-diabetic CGM trace are repeatable and physiologically meaningful. Specifically, clinicians and researchers need to know how many days of wear establish a reliable glycemic baseline, which metrics most strongly reflect insulin resistance (the metabolic defect that precedes dysglycemia by years^12^), whether CGM uncovers early metabolic risk that a normal HbA1c (<5.7%) misses,^13^ and which aspects of post-meal excursions and food responses actually track cardiometabolic health.

Answering these questions requires observing continuous glucose, meal composition, and clinical biomarkers in the same individuals at scale, whereas existing studies have relied largely on single wear periods or smaller clinic-based cohorts.^14–16^ Here, by linking free-living CGM recordings, logged meals, and commercial laboratory results across 62,122 consenting adults on a digital-health platform, we show that CGM captures insulin-resistance signal that HbA1c misses and identify which glucose spike features can guide preventive care.

## Results

### A linked, multimodal cohort of free-living adults

Between 2022 and 2025, 62,122 adults consented to share data from the Levels platform: 49,876 contributed CGM recordings, 53,595 logged behavior, 12,307 shared laboratory results, and 10,924 contributed all three streams (Fig. 1a,b). Because each stream was opt-in, sensor wear was episodic across a median consented span of 772.1 days, with a median of 42 days with CGM data per participant. Participants were predominantly in their 30s to 50s (74.6%), and 53.7% were female (Fig. 1c). By their highest recorded HbA1c or fasting glucose, 63.2% of participants with glycemic results fell in the normal range, 33.4% in the prediabetes range, and 3.4% in the diabetes range (Fig. 1d). Excluding anyone with a diabetes-range laboratory value yielded a primary laboratory cohort of 7,930 adults without diabetes (Fig. 1b; Table 1).

**Fig. 1.**
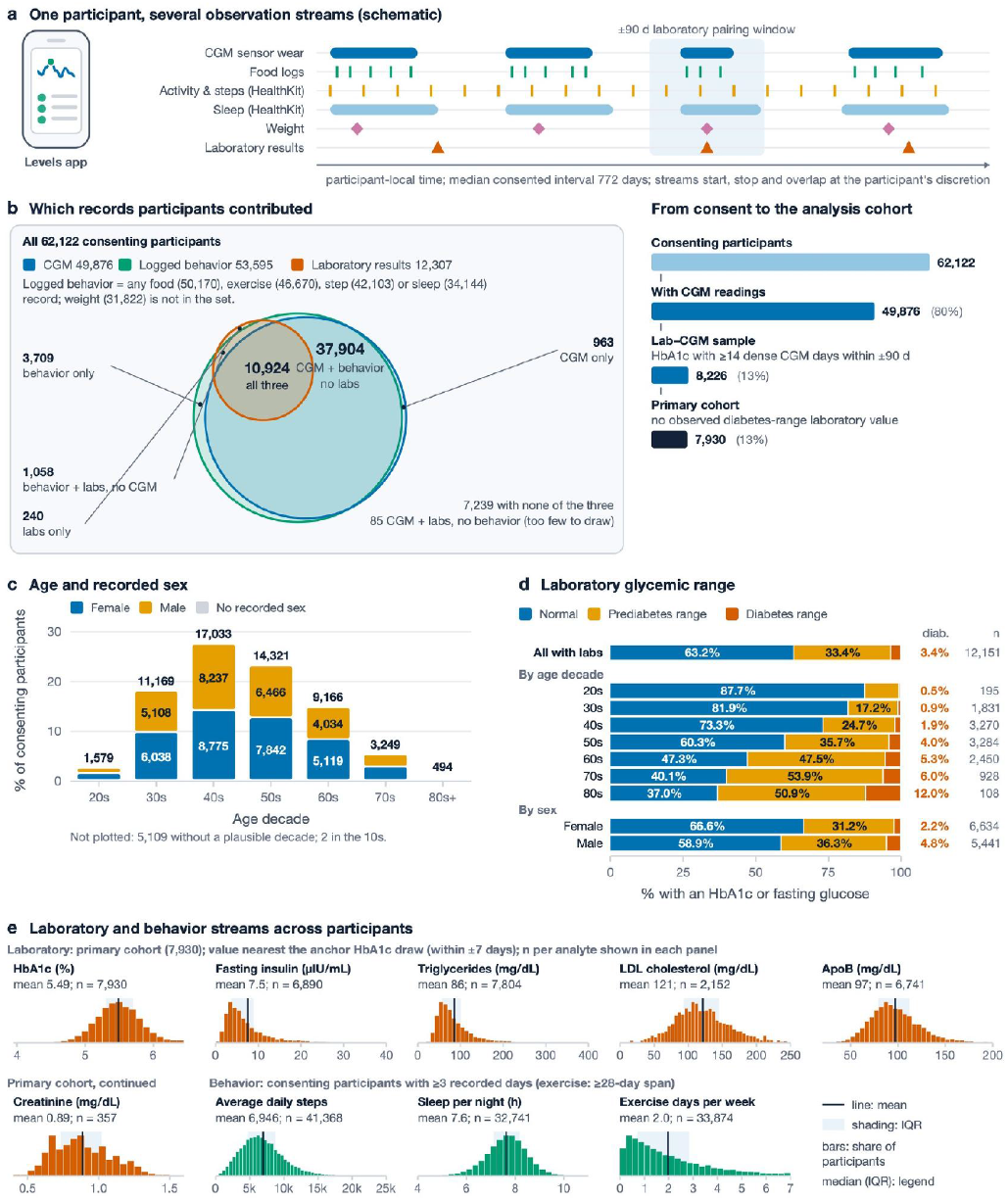
| Cohort and linked data streams. **a**, Schematic of one participant’s observation streams and the ±90-day window within which CGM is paired with a laboratory draw; the span shown is the median consented interval (772 days), not continuous sensor wear. **b**, Left, overlap of CGM (49,876 participants), logged behavior (53,595; any food, exercise, step, or sleep record), and laboratory results (12,307); 10,924 contributed all three. Right, flow from 62,122 consenting participants to 49,876 with CGM, 8,226 with an HbA_1c_ and at least 14 dense CGM days within ±90 days, and 7,930 in the primary cohort without an observed diabetes-range laboratory value. **c**, Age decade and recorded sex of consenting participants. **d**, ADA range of each participant’s highest accepted HbA_1c_ or fasting glucose (12,151 participants): normal 63.2%, prediabetes range 33.4%, diabetes range 3.4%. Single measurements, not diagnoses. **e**, Distributions across participants (line, mean; shading, IQR). Primary cohort, values nearest the anchor HbA_1c_ draw (within ±7 days), median (IQR): HbA_1c_ 5.5% (5.3–5.7); fasting insulin 5.6 µIU/mL (3.6–8.9); triglycerides 73 mg/dL (57–101); LDL cholesterol 117 mg/dL (93–145; 2,152 participants); ApoB 94 mg/dL (79–112); creatinine 0.86 mg/dL (0.73–1.02; 357 participants). Consenting participants with enough records: daily steps 6,578 (4,681–8,744); sleep 7.7 h per night (7.1–8.2); exercise 1.5 days per week (0.7–2.8). ADA, American Diabetes Association; ApoB, apolipoprotein B; CGM, continuous glucose monitoring; IQR, interquartile range; LDL, low-density lipoprotein.

**Table 1.**
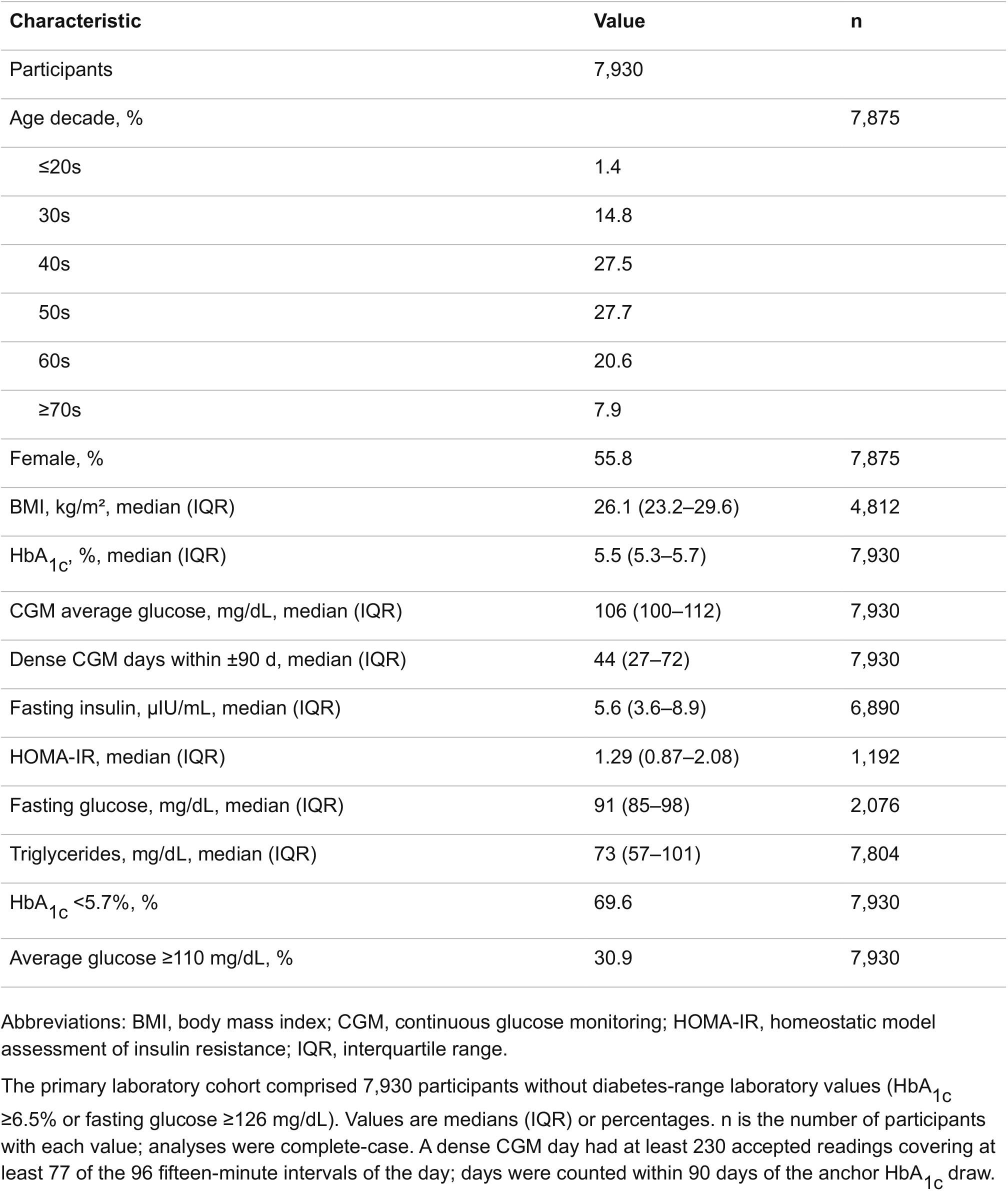
| Characteristics of the primary laboratory cohort.

### One week of CGM wear captures a reliable glycemic baseline

Across 36,454 participants with at least two separate weeks of wear, average glucose was the most repeatable CGM measure from the first week to the second (intraclass correlation, ICC 0.747), outperforming the standard deviation (SD), the mean of daily differences (MODD), and the CV (Fig. 2a,b). Across two wear windows separated by 14 days, reliability for average glucose plateaued at approximately 7 days of wear (ICC 0.728; Fig. 2c), with variability metrics following similar trajectories. Agreement between 7-day recordings declined gradually as the interval between wear periods grew, from 0.832 for back-to-back weeks to 0.628 at 36 to 90 days and 0.561 beyond 90 days (Fig. 2b), consistent with a likely mixture of average glucose tracking a slow-moving metabolic state and differing sensor bias drift.

**Fig. 2.**
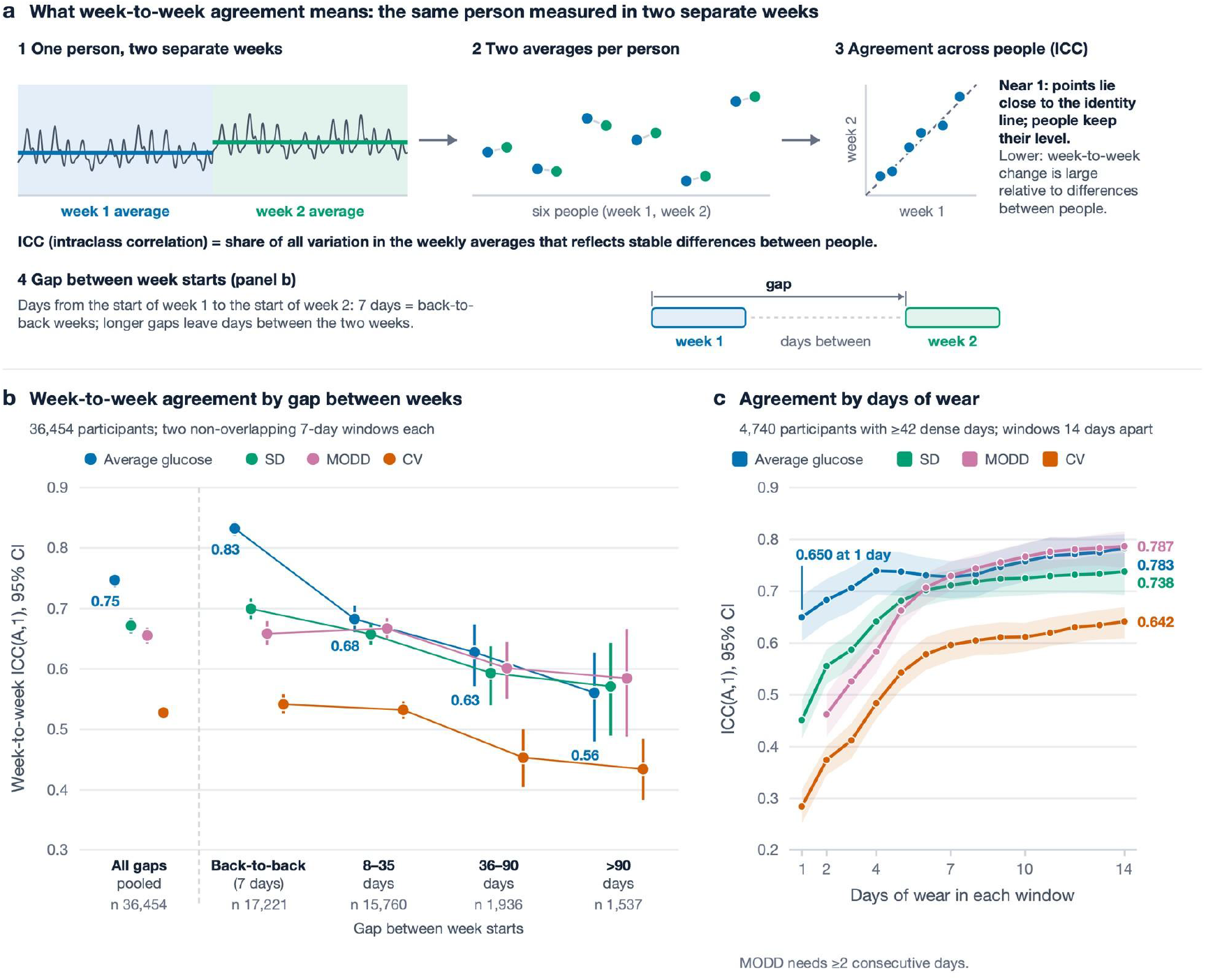
| How long to wear a sensor. **a**, What week-to-week agreement measures (schematic): the intraclass correlation (ICC) is the share of variation in weekly averages that reflects stable differences between people; the gap is the number of days between the starts of the two weeks. **b**, Week-to-week ICC by CGM measure, pooled and by gap between week starts, in 36,454 participants; values under the markers are for average glucose (0.747 pooled; 0.832 for back-to-back weeks; 0.561 for weeks more than 90 days apart). Part of the fall over longer gaps may reflect real change in metabolic state. **c**, ICC by days of wear in two windows 14 days apart, in 4,740 participants with at least 42 dense days: average glucose 0.650 at 1 day, 0.739 at 4 days, and 0.783 at 14 days. MODD needs at least 2 consecutive days. Participants without an observed diabetes-range laboratory value; windows of 7 consecutive dense days. Error bars and bands, 95% participant-bootstrap confidence intervals. CGM, continuous glucose monitoring; CV, coefficient of variation; ICC, intraclass correlation coefficient, ICC(A,1); MODD, mean of daily differences; SD, standard deviation.

### Average glucose most reliably tracks insulin resistance and metabolic health

In the primary cohort, each SD (9.1 mg/dL) of higher average glucose was associated with 28.3% higher fasting insulin (95% CI 26.2% to 30.4%; 6,890 participants) and 30.0% higher HOMA-IR, with the prevalence of HOMA-IR ≥2.5 rising nearly fivefold from the lowest to the highest quartile of average glucose (Fig. 3a,d). Controlling for BMI softened but preserved this association (16.0% higher fasting insulin per SD in 4,257 participants; Fig. 3b). Assessed individually, variability metrics (MODD, SD, and MAGE), CGM-detected spikes (frequency, height, and width), and logged-meal responses (2-hour incremental area under the curve, iAUC) were also associated with higher fasting insulin, though less strongly than average glucose, whereas the CV was not (Fig. 3d).

**Fig. 3.**
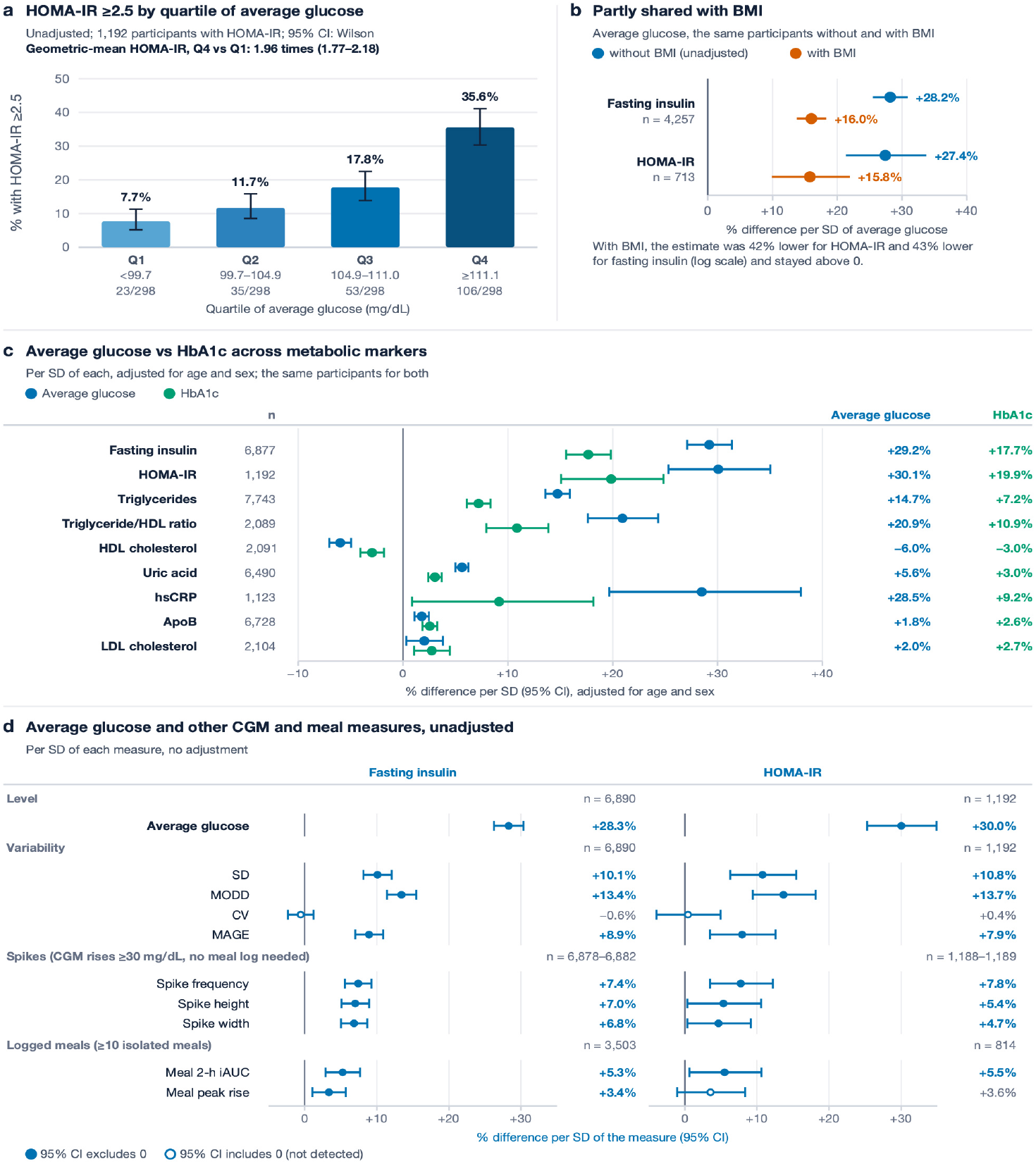
| Average glucose tracks insulin resistance better than other CGM markers. **a**, Share with HOMA-IR ≥2.5 by quartile of average glucose (1,192 with HOMA-IR; Wilson 95% CI): from 7.7% (lowest quartile) to 35.6% (highest); geometric-mean HOMA-IR 1.96 times as high in the highest as in the lowest quartile (95% CI 1.77 to 2.18). **b**, Average glucose without and with adjustment for BMI in the same participants: per SD, fasting insulin (4,257 participants) +28.2% and +16.0%; HOMA-IR (713) +27.4% and +15.8%. **c**, Percentage difference in each marker per SD of average glucose (blue) and per SD of HbA1c (green), in the same participants, each model adjusted for age decade and sex: fasting insulin +29.2% vs +17.7% (6,877); HOMA-IR +30.1% vs +19.9% (1,192); triglycerides +14.7% vs +7.2% (7,743); triglyceride/HDL ratio +20.9% vs +10.9% (2,089); HDL cholesterol −6.0% vs −3.0% (2,091); uric acid +5.6% vs +3.0% (6,490); hsCRP +28.5% vs +9.2% (1,123); ApoB +1.8% vs +2.6% (6,728); LDL cholesterol +2.0% vs +2.7% (2,104). Average glucose showed the larger association for every marker except ApoB and LDL cholesterol, for which both were small. **d**, Percentage difference in fasting insulin (6,890 participants) and HOMA-IR (1,192) per SD of each measure, unadjusted; average glucose +28.3% and +30.0%; spike width (minutes that glucose stays above half the rise of each spike): fasting insulin +6.8% (95% CI 5.0% to 8.7%), HOMA-IR +4.7% (95% CI 0.4% to 9.2%). Logged-meal responses are means over at least 10 isolated logged meals. Every other measure was less strongly related than average glucose for both markers. Primary cohort without diabetes-range laboratory values (7,930 participants). Filled markers, 95% CI excludes 0; open markers, CI includes 0 (not detected). Error bars, 95% confidence intervals (Wilson in a; heteroskedasticity-consistent in b–d). ApoB, apolipoprotein B; BMI, body mass index; CGM, continuous glucose monitoring; CV, coefficient of variation; HOMA-IR, homeostatic model assessment of insulin resistance; hsCRP, high-sensitivity C-reactive protein; iAUC, incremental area under the curve; LDL, low-density lipoprotein; MAGE, mean amplitude of glycemic excursions; MODD, mean of daily differences; SD, standard deviation.

Average glucose also tracked broader cardiometabolic biomarkers more closely than HbA1c (Fig. 3c; Supplementary Table 1a). Each SD of average glucose showed a stronger association than each SD of HbA1c (adjusted for age and sex) and explained more variance across triglycerides (14.7%, 95% CI 13.6% to 15.9%, vs 7.2%), the triglyceride-to-HDL ratio, HDL cholesterol, uric acid, and high-sensitivity C-reactive protein (hsCRP). Neither metric explained more than 1.5% of the variance in apolipoprotein B (ApoB) or LDL cholesterol, consistent with average glucose reflecting insulin resistance rather than atherogenic lipoprotein load.

### Spike width rather than peak height carries independent metabolic risk signal

Because people with higher average glucose also experience larger and more frequent excursions, we controlled for average glucose to isolate which postprandial features add independent information (Fig. 4a). Counterintuitively, at the same average glucose, the frequency and height of CGM-detected spikes and the iAUC of logged meals did not indicate higher insulin resistance; they were instead associated with slightly lower fasting insulin (for example, 5.1% lower per SD of spike height; Fig. 4b).

**Fig. 4.**
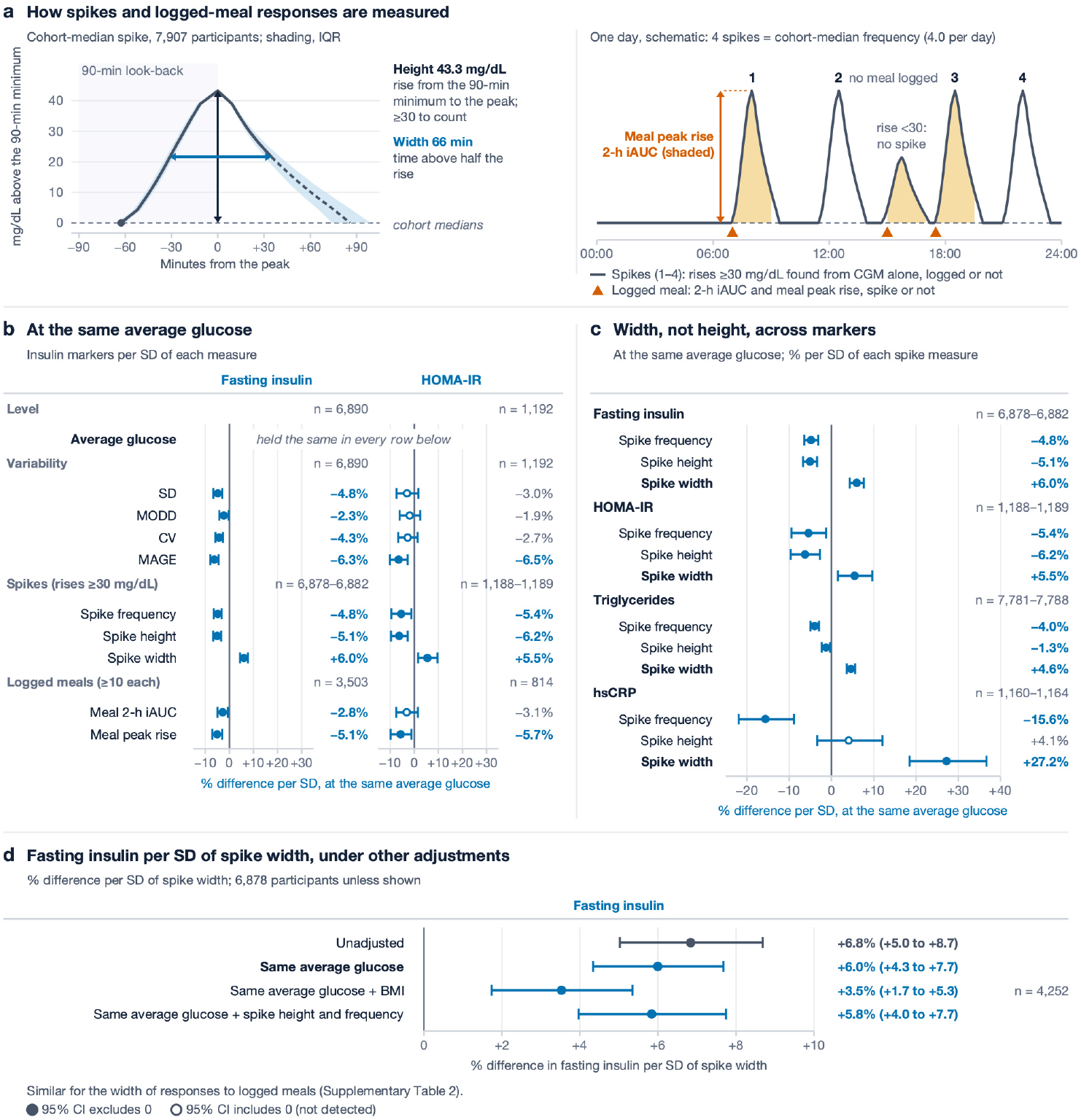
| Spike width, not height, tracks insulin resistance and metabolic markers at the same average glucose. **a**, Left, the cohort-median spike (7,907 participants; 1,128,300 spikes that fell at least halfway back): for each tenth of the rise, the median across participants of their average time to reach it (shading, IQR; dashed below half the rise on the way down, from spikes that fell that far). A spike is any CGM rise of at least 30 mg/dL above the lowest value of the preceding 90 min, detected without meal logs. Height, rise from that minimum to the peak; width, time above half the rise. Cohort medians (IQR) of participant averages (7,907–7,914 participants): height 43.3 (41.0–46.0) mg/dL, width 66 (60–73) min, frequency 4.0 (2.9–5.2) per day. Right, schematic day built from the same shape at the cohort-median frequency; logged-meal responses (2-h iAUC, shaded; meal peak rise) are measured only after logged isolated meals, whether or not a spike follows. **b**, Percentage difference per SD of each measure at the same average glucose (fasting insulin, 6,890 participants; HOMA-IR, 1,192). Only spike width had a 95% CI above 0 for both markers: fasting insulin +6.0% (95% CI 4.3 to 7.7), HOMA-IR +5.5% (1.5 to 9.6); spike height −5.1% and −6.2%. **c**, The same comparison for triglycerides and hsCRP (fasting insulin and HOMA-IR as in b). Width: triglycerides +4.6% (95% CI 3.6 to 5.6; 7,781 participants), hsCRP +27.2% (18.5 to 36.7; 1,160). Height: triglycerides −1.3%, hsCRP +4.1% (not detected); frequency: −4.0% and −15.6%. **d**, Fasting insulin per SD of spike width under other adjustments: unadjusted +6.8%; same average glucose +6.0%; plus BMI +3.5% (95% CI 1.7 to 5.3; 4,252 participants with BMI); plus spike height and frequency +5.8%. The width of responses to logged meals gave similar results (Supplementary Table 2). OLS of the log marker, heteroskedasticity-consistent 95% CIs. Filled markers, 95% CI excludes 0; open, not detected. hsCRP, high-sensitivity C-reactive protein; iAUC, incremental area under the curve; CV, MAGE, and MODD as in Fig. 3.

By contrast, spike width, defined as the duration glucose remained above half of its rise (cohort median 66 minutes), maintained a positive association. At the same average glucose, each SD of wider spikes (about 11 minutes longer) was associated with 6.0% higher fasting insulin (95% CI 4.3% to 7.7%; 6,878 participants), alongside higher HOMA-IR (Fig. 4b), triglycerides, and hsCRP (Fig. 4c). This association with fasting insulin held without adjustment, after further adjustment for BMI or for spike height and frequency (Fig. 4d), and when width was measured from isolated logged meals (Supplementary Table 2). Spike height therefore mainly reflected baseline average glucose, whereas wider spikes marked elevated fasting insulin, triglycerides, and hsCRP at a given average glucose.

### CGM uncovers elevated glucose and insulin that a normal HbA1c does not detect

Of the 7,930 participants in the primary cohort, classified by average glucose (≥110 vs <110 mg/dL) and HbA1c (≥5.7% vs <5.7%), 4,212 (53.1%) were normal on both measures, 1,146 (14.5%) were elevated on both, and 2,572 (32.4%) were discordant: 1,304 (16.4%) had elevated average glucose with a normal HbA1c, and 1,268 (16.0%) had an elevated HbA1c with normal average glucose (Fig. 5d). Nearly one in four participants with a normal HbA1c (23.6%) therefore had elevated average glucose. Across all 8,226 participants with paired measurements, including 296 with diabetes-range values, our data reproduced the slope of the published glucose management indicator (GMI) calibration,^17^ though with a different intercept (Supplementary Fig. 1). Within the normal HbA1c range (<5.7%), however, the relationship was nearly flat: each 0.1-point HbA1c step corresponded to only 0.64 mg/dL higher average glucose, one-third of the 1.95 mg/dL slope within the prediabetes range (5.7–6.4%; Fig. 5a), with a weaker correlation (r = 0.17 vs 0.34; Fig. 5b).

**Fig. 5.**
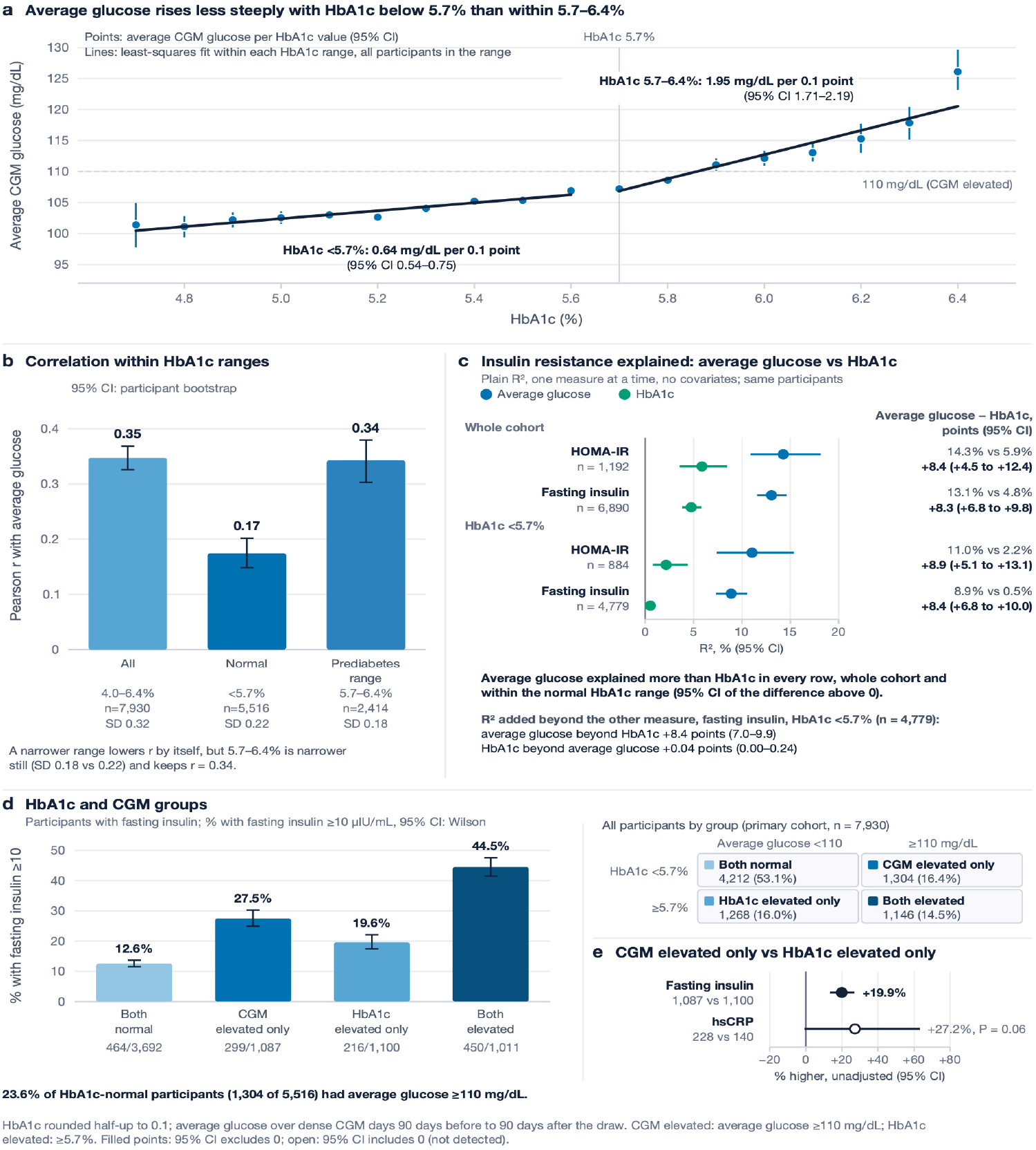
| Average CGM glucose tracks insulin resistance more closely than HbA1c. **a**, Average glucose at each HbA1c value with at least 30 participants (18 values; 95% CI), with least-squares fits within each range: 0.64 mg/dL per 0.1 HbA1c point below 5.7% (95% CI 0.54 to 0.75) and 1.95 within 5.7–6.4% (1.71 to 2.19); difference 1.31 (1.04 to 1.57; P < 0.001). Dashed line, 110 mg/dL threshold used in d. **b**, Pearson correlation of HbA1c with average glucose: 4.0–6.4%, r = 0.35 (7,930 participants); <5.7%, r = 0.17 (5,516); 5.7–6.4%, r = 0.34 (2,414); participant-bootstrap 95% CI. **c**, Variance in log fasting insulin and log HOMA-IR explained (R²) by average glucose vs by HbA1c alone, in the same participants. Whole cohort: fasting insulin (6,890) 13.1% vs 4.8%; HOMA-IR (1,192) 14.3% vs 5.9%. HbA1c <5.7%: fasting insulin (4,779) 8.9% vs 0.5%; HOMA-IR (884) 11.0% vs 2.2%; the 95% CI of the difference excluded 0 in every row. Within HbA1c <5.7%, average glucose added 8.4 percentage points of R² for fasting insulin beyond HbA1c (95% CI 7.0 to 9.9); HbA1c added 0.04 (0.00 to 0.24) beyond average glucose. **d**, The 2 × 2 discordance groups of HbA1c × CGM in the primary cohort: both normal 4,212 (53.1%); CGM elevated only 1,304 (16.4%); HbA1c elevated only 1,268 (16.0%); both elevated 1,146 (14.5%). Bars, share with fasting insulin ≥10 µIU/mL (Wilson 95% CI): both normal 12.6% (464 of 3,692); CGM elevated only 27.5% (299 of 1,087); HbA1c elevated only 19.6% (216 of 1,100); both elevated 44.5% (450 of 1,011). **e**, Fasting insulin was 19.9% higher (95% CI 13.3% to 26.8%; 1,087 vs 1,100 participants) and hsCRP 27.2% higher (95% CI −0.8% to 63.2%; P = 0.06; 228 vs 140; open marker, CI includes 0) in the CGM-elevated-only than in the HbA1c-elevated-only group (ratio of geometric means). Primary cohort without diabetes-range laboratory values (7,930 participants). CGM elevated, average glucose ≥110 mg/dL; HbA1c elevated, ≥5.7%. Error bars, 95% confidence intervals. CGM, continuous glucose monitoring; HOMA-IR, homeostatic model assessment of insulin resistance; hsCRP, high-sensitivity C-reactive protein.

Although CGM–HbA1c discordance has been described previously,^18^ paired biomarkers revealed that CGM uncovered elevated fasting insulin substantially better than HbA1c (Fig. 5c–e; Supplementary Table 3). Average glucose explained nearly three times more variance in fasting insulin than HbA1c across the primary cohort (R² 0.131 vs 0.048) and 8.9% vs 0.5% within the normal HbA1c range, where HbA1c added almost nothing once average glucose was known (0.04 percentage points; Fig. 5c). Among participants with a normal HbA1c, fasting hyperinsulinemia (≥10 µIU/mL) was more than twice as common at average glucose ≥110 mg/dL as below (27.5% vs 12.6%; prevalence ratio 2.2, 95% CI 1.9 to 2.5; 4,779 participants; Fig. 5d). Directly comparing the two discordant groups, participants with elevated average glucose and a normal HbA1c had 19.9% higher fasting insulin (P < 0.001), more frequent hyperinsulinemia (27.5% vs 19.6%), higher HOMA-IR and triglycerides (Supplementary Table 1b), and a trend toward higher hsCRP (27.2% higher; P = 0.06; Fig. 5d,e) than those with an elevated HbA1c and normal average glucose.

Both HbA1c and fasting insulin were repeatable across draws 14 to 90 days apart (ICC 0.72 in 607 pairs and 0.71 in 449 pairs; Supplementary Table 4), confirming that the weak resolution of HbA1c in the normal range did not reflect assay noise. Participants with elevated average glucose and a normal HbA1c were as young as those normal on both measures (53% vs 51% under 50 years), whereas only 27% of those with an elevated HbA1c alone were under 50. Adjusted for age and sex, the CGM-elevated group was also heavier (BMI 2.1 kg/m² higher, 95% CI 1.7 to 2.5), less active, and slept less than participants normal on both measures (Supplementary Fig. 2), pointing to particular utility for CGM screening in younger adults.

### Repeated meals capture personal food responses, but carbohydrate sensitivity does not track insulin resistance

Pairing CGM with logged meals allowed us to estimate how many repeated meals are needed to characterize a person’s response to a specific food. Two isolated meals of the same labeled food eaten by the same person agreed only modestly (r = 0.42, 95% CI 0.40 to 0.44; 10,486 participants; Fig. 6a,b), matching the agreement of duplicate meals served a week apart in inpatient feeding trials (r = 0.45 and 0.46 across two sensor models).^19^ However, averaging five isolated meals of the same food across sets separated by a median of 29 days yielded reliable agreement (r = 0.72; 1,131 participants; Fig. 6b), demonstrating that multi-meal averaging captures reproducible personal food responses in daily life.

**Fig. 6.**
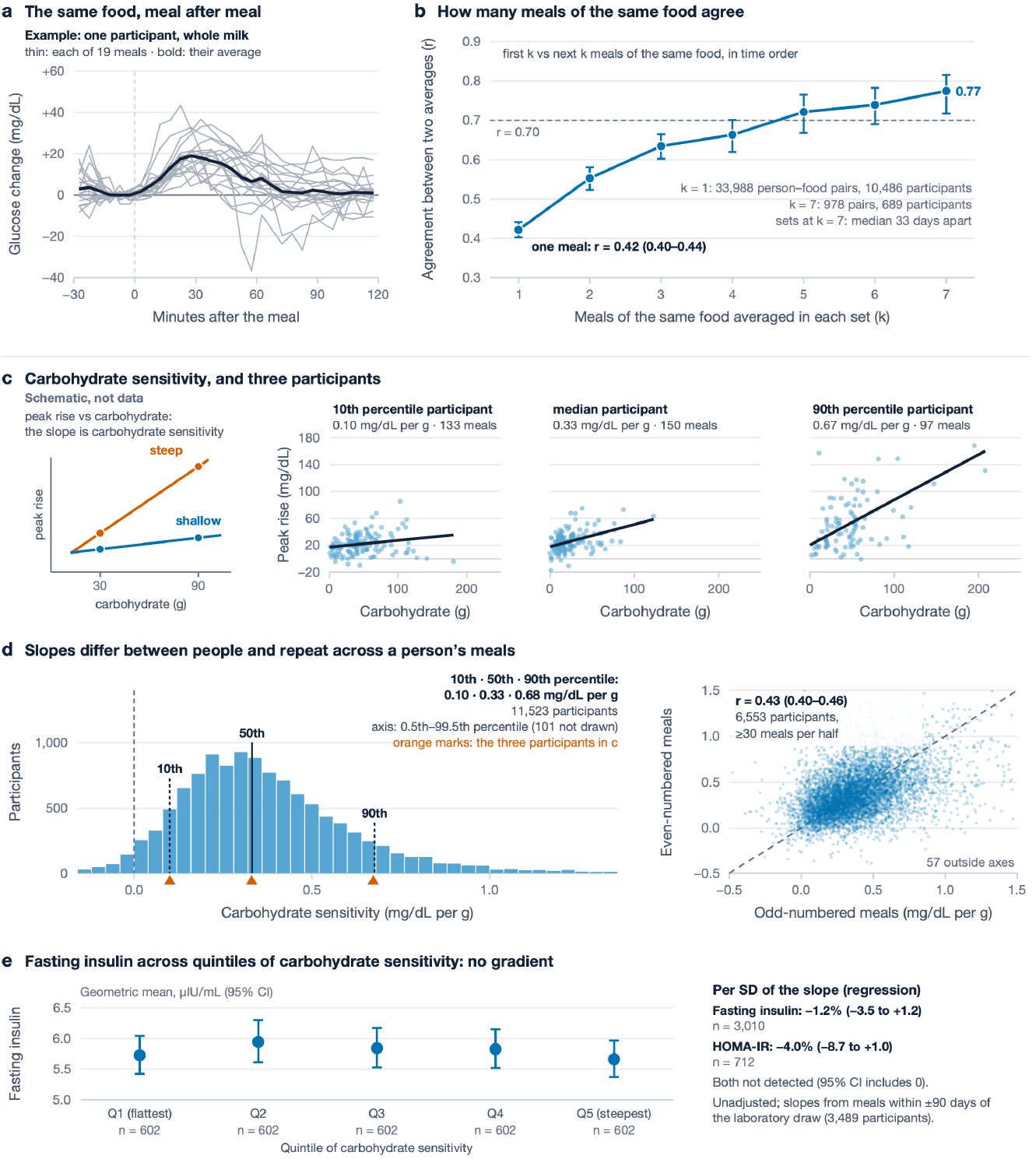
| Reliability of food responses and carbohydrate sensitivity. **a**, Example of one de-identified participant: 19 meals of whole milk (thin lines, glucose change from the premeal level) and their average (bold). **b**, Agreement (Pearson r) between the averages of the first k and the next k meals of the same food in the same person: 0.42 for one meal (33,988 person–food pairs, 10,486 participants); above 0.70 from five meals (0.72; 1,131 participants; sets a median 29 days apart); 0.77 for seven meals (689 participants; a median 33 days apart). **c**, Carbohydrate sensitivity, the slope of meal peak rise on carbohydrate at the same fiber, fat, and protein: schematic and three de-identified participants nearest the 10th, 50th, and 90th percentile slopes. **d**, Distribution of slopes in 11,523 participants (10th and 90th percentiles, 0.10 and 0.68 mg/dL per g) and agreement between odd– and even-numbered meals (r = 0.43; 6,553 participants). **e**, Geometric-mean fasting insulin by quintile of carbohydrate sensitivity (602 participants per quintile; slopes recomputed from meals within ±90 days of the laboratory draw): 5.7 to 5.9 µIU/mL, with no trend (−0.4% per quintile; 95% CI −2.1% to 1.3%). Per SD of the slope, fasting insulin −1.2% (95% CI −3.5% to 1.2%; 3,010 participants) and HOMA-IR −4.0% (95% CI −8.7% to 1.0%; 712); neither association was detected. Unadjusted. Self-logged meals. Error bars, 95% confidence intervals. HOMA-IR, homeostatic model assessment of insulin resistance.

Across each participant’s logged meals (≥30 meals), carbohydrate sensitivity, defined as the rise in peak glucose per gram of carbohydrate holding fiber, fat, and protein constant, varied sevenfold between the 10th and 90th percentiles (11,523 participants) and was moderately repeatable across odd– and even-numbered meals (r = 0.43, 95% CI 0.40 to 0.46; Fig. 6c,d). Although often targeted by personalized nutrition advice,^20,21^ carbohydrate sensitivity did not track insulin resistance: geometric mean fasting insulin was flat across quintiles of carbohydrate sensitivity (trend −0.4% per quintile, 95% CI −2.1% to 1.3%; 3,010 participants), and neither fasting insulin nor HOMA-IR varied per SD of the slope (Fig. 6e).

## Discussion

The rapid uptake of over-the-counter CGMs by adults without diabetes has prompted debate over whether their traces offer meaningful clinical information.^7,8,10,11^ Across 62,122 free-living adults, including 7,930 with linked laboratory results, CGM captured cardiometabolic risk signal in adults without diabetes, particularly within the normal HbA1c range where HbA1c is least informative. Our findings also reframe post-meal interpretation: single meals and peak spike height are weak indicators of insulin resistance, whereas multi-meal averages are reproducible and spike width carries independent physiological signal.

For clinicians, these results suggest a practical framework for preventive care (Box 1). First, approximately 7 days of wear captures a reliable glycemic baseline, and average glucose remains reasonably stable across wear periods up to 90 days apart (ICC 0.628 at 36 to 90 days), a timescale comparable to the three-month window of HbA1c. Second, because CGM measures interstitial glucose directly, it avoids non-glycemic influences on HbA1c, which depends on red-cell lifespan and glycation rates^22^ and rises with age independently of glucose^23^ (Supplementary Fig. 3). Consequently, a fixed 5.7% HbA1c threshold may under-detect early insulin resistance in younger adults while over-calling dysglycemia in older adults. While HbA1c remains an established predictor of long-term vascular outcomes,^24^ multi-week average glucose is a complementary measure that is more sensitive to early insulin resistance, particularly in younger adults with normal HbA1c.

Third, our findings clarify the physiology of post-meal spikes. Considered in isolation, individuals with taller or more frequent spikes have higher fasting insulin simply because excursion amplitude rises with average glucose. At the same average glucose, however, taller or more frequent spikes were associated with slightly lower fasting insulin, likely because larger daytime excursions mathematically require a lower inter-meal glucose floor to maintain the same mean. By contrast, wider spikes were associated with higher fasting insulin, HOMA-IR, triglycerides, and hsCRP at the same average glucose. This mirrors oral glucose tolerance testing, where a delayed glucose peak marks reduced β-cell function and prediabetes risk,^25^ and aligns with clinic-based CGM studies linking higher glucose autocorrelation (prolonged elevations) to hepatic and vascular risk independently of mean glucose.^16^ In a remote multimodal cohort, the time to clear half of a glucose spike was longer in type 2 diabetes than in normoglycemia.^26^ Because fasting insulin and HOMA-IR primarily reflect hepatic insulin resistance, whereas postprandial glucose clearance depends on incretin-potentiated insulin secretion and skeletal muscle uptake,^27,28^ the association between spike width and fasting insulin (+6.0% per SD) likely understates its value as a dynamic marker of peripheral insulin sensitivity.^28^ Prospective interventions can now test whether narrowing spike width, rather than blunting peak height, improves cardiometabolic risk.

These results also inform personalized nutrition, which has largely aimed to minimize acute postprandial glucose peaks.^20,21^ Although responses to standardized carbohydrate loads correlate with insulin resistance and β-cell function in controlled clinical settings,^27^ free-living carbohydrate sensitivity did not track insulin resistance here, challenging its use as a standalone proxy for metabolic health in daily life, though we cannot rule out that minimizing spikes may provide benefits not captured by standard biomarkers. Other features of the postprandial curve may also matter: the dip below baseline 2–3 h after a meal, rather than peak height, has been linked to later hunger and energy intake,^29,30^ outcomes that this analysis, limited to CGM traces as markers of underlying metabolic risk, could not capture. Furthermore, because single meals agreed only modestly (r = 0.42), dietary guidance should rely on averaging at least five repeated meals (r = 0.72).

Similarly, these principles set a benchmark for AI foundation models trained on CGM traces, one of which recently outperformed HbA1c in predicting incident diabetes and cardiovascular death over a median of 11 years.^5^ Our data suggest that average glucose and spike width carry much of the signal such models could exploit; new algorithms should therefore demonstrate incremental value beyond multi-week average glucose and spike width across independent wear periods.

While free-living observation captures physiology in daily life, this cross-sectional study has important limitations. Participants were self-selected, fee-paying users of a commercial platform, meal logging was self-reported, and sensor models and interface workflows evolved over time, although associations were consistent across anchor years (Supplementary Table 3). Because medical records, race and ethnicity, and medication use (including metformin or glucagon-like peptide-1 receptor agonists) were unavailable, diabetes exclusion relied on laboratory values alone. Fasting insulin and HOMA-IR are single-time-point surrogates of primarily hepatic insulin resistance, fasting status was self-reported, and commercial insulin assays are not standardized. Finally, pairing blood draws with CGM worn up to 90 days apart introduces temporal noise that would attenuate CGM associations; because HbA1c and fasting insulin were drawn together, this design would, if anything, favor HbA1c in head-to-head comparisons. Because CGM and HbA1c were compared in the same participants against the same laboratory draws, these limitations are unlikely to explain why average glucose tracked insulin more closely than HbA1c.

As human health becomes continuously observable, consumer digital-health platforms offer a high-resolution window into early metabolic physiology. In adults without diabetes, CGM uncovers elevated glucose and insulin resistance that a normal HbA1c misses, and provides actionable postprandial signal when clinical and consumer interpretation shifts from the height of a single spike to multi-week average glucose and spike width.

### Box 1 | A clinical framework for CGM in adults without diabetes

**Why use CGM when HbA_1c_ is normal (<5.7%)?** About one in four adults with a normal HbA_1c_ have an average glucose of 110 mg/dL or higher, and in them elevated fasting insulin (≥10 µIU/mL) is twice as common. CGM may be most informative in younger adults, in whom HbA_1c_ is lower at the same glucose.

**How long to wear a sensor.** Aim for at least 7 days to establish a reliable baseline. Because a person’s glucose changes over months (agreement falls from 0.832 for back-to-back weeks to 0.561 for weeks more than 90 days apart), periodic multi-week wear is more informative than a single short recording.

**Primary target: average glucose.** Prioritize multi-week average glucose, which explains nearly three times as much variation in fasting insulin as HbA_1c_ (R² 0.131 vs 0.048). A persistent average glucose of 110 mg/dL or higher may warrant checking fasting insulin and triglycerides.

**How to read spikes: width, not height.** At the same average glucose, spike width (how long glucose stays above half of its rise) tracks fasting insulin, triglycerides, and hsCRP, whereas peak height and frequency do not and may reflect a lower baseline between meals.

**How to guide food testing.** Counsel patients not to judge a food from a single meal (r = 0.42); averaging about five meals of the same food (r = 0.72) identifies a reliable personal response.

## Methods

### Study design and participants

This cross-sectional study analyzed a de-identified research export from the Levels digital-health platform, collected between 2022 and 2025 under a decentralized, non-interventional protocol (LH-001) sponsored by Levels Health, Inc. and approved by the Advarra Institutional Review Board (Pro00058897). Adult US members provided electronic informed consent, including HIPAA authorization. Exclusion criteria at screening were exogenous insulin use, type 1 diabetes, or type 2 diabetes treated with medications other than metformin or pioglitazone. Participants paid for platform membership, purchased commercially available CGM sensors, selected which data streams to share, and received no research compensation. Clinical diagnoses, medication records, race and ethnicity, and specific sensor models were not included in the research export. Reporting follows the STROBE guideline.^31^

### Platform and data streams

Users joined the direct-to-consumer platform voluntarily and opted separately into research data sharing. Over the study period, supported CGM brands, device generations, app interfaces, and logging workflows evolved. CGM traces were collected via platform-connected sensors; food, exercise, alcohol, and body weight were logged by users; and step counts and sleep were imported when users enabled Apple HealthKit integration. Laboratory results originated predominantly from commercial clinical laboratories (Labcorp and Quest) via direct platform integrations, with a minority self-reported; draws were not at study-defined intervals. Each participant’s local calendar day was defined using a single fixed UTC offset derived from the modal device-clock offset across their food logs (or the cohort mode when food logs were absent); week-to-week reliability analyses required a participant-specific food-log offset. Food, exercise, and alcohol entries were assigned to the date of their start timestamp, sleep to its wake date, and steps and weight to their recorded date.

### CGM measures

Accepted CGM readings (20 to 500 mg/dL) were averaged within 15-minute intervals of each local calendar day. Metrics were computed on dense days, defined by at least 230 accepted readings covering at least 77 of the 96 daily intervals. Interior gaps of one or two intervals within a contiguous run of dense days were linearly interpolated; boundary gaps were not interpolated, and dense-day eligibility used observed intervals only. Average glucose was the mean of the interval values. Summary metrics (Supplementary Table 5) included average glucose, SD, CV (100 × SD/mean), MODD (mean absolute difference between matching clock intervals on consecutive dense days), and MAGE. MAGE was computed across all accepted dense-day readings per participant using the moving-average algorithm of Fernandes et al.^32^ in the R package iglu^33^ (version 4.2.2, default settings: 5-minute grid interpolation across gaps ≤45 min, segmentation at gaps >3 h, peak/nadir detection via 5-point and 32-point moving-average crossings, and duration-weighted averaging of ascending and descending excursions exceeding 1 segment SD).

Spikes were detected directly from uninterpolated 5-minute CGM intervals without meal logs. Adapting the definition of Carletti et al.,^26^ a spike was defined as a local maximum at least 30 mg/dL above the minimum of the preceding 90 minutes; when two peaks occurred within 30 minutes, only the higher peak was retained. Days were eligible for spike analysis if at least 16 hours (192 five-minute intervals with complete preceding 90-minute and following 5-minute coverage) were observable. Spikes were characterized by frequency (mean spikes per eligible day), height (rise from the 90-minute minimum to peak), and width (minutes spent above half the rise from upslope to downslope, evaluated on spikes returning to half-height within 6 hours and before a subsequent peak, missing interval, or midnight; Fig. 4a). Participant-level spike metrics required at least 5 eligible days and, for width, at least 5 resolved spikes.

### Logged meals

Food logs entered within 15 minutes of one another were merged into a single meal timed at the initial entry; multi-entry meals spanning more than 15 minutes or lacking nutrient entries were excluded. An isolated meal required no other logged food within 2 hours before or after it (unlogged intake cannot be excluded) and complete uninterpolated 5-minute CGM coverage from 15 minutes pre-meal through 2 hours post-meal. Using mean glucose over the 15 minutes pre-meal as baseline, we computed 2-hour iAUC (area above baseline), meal peak rise (maximum 2-hour glucose minus baseline), and, for meals rising at least 20 mg/dL, response width (minutes above half-rise within 180 minutes, averaged across ≥10 qualifying meals per participant within ±90 days of the anchor draw; Supplementary Table 2).

Meal macronutrients were summed across constituent items; if any item had a missing or implausible value for a given nutrient, that nutrient was treated as missing (never imputed as zero). Carbohydrate sensitivity was estimated per participant as the ordinary least-squares slope of meal peak rise on carbohydrate (g), controlling for fiber, fat, and protein across at least 30 complete meals with a carbohydrate SD ≥5 g (Supplementary Table 5). Repeated meals of the same food were isolated single-item meals matched by normalized food name or barcode.

### Laboratory pairing

For each participant, the anchor draw was the earliest HbA1c measurement accompanied by at least 14 dense CGM days within ±90 days. Companion analytes were taken from the draw nearest the anchor within ±7 days, and CGM metrics were aggregated across all dense days within the ±90-day window. Fasting status was self-reported. Analyte units were standardized using explicit conversion rules and analytic validity ranges, excluding pmol/L insulin values lacking assay-specific conversion factors (Supplementary Table 6). Same-day duplicate labs were collapsed to their median (excluding anchor dates with same-day HbA1c discrepancies >0.3 percentage points), with HbA1c rounded half-up to 0.1%. HOMA-IR (fasting glucose [mg/dL] × fasting insulin [µIU/mL] / 405)^34^ and the triglyceride-to-HDL ratio required same-day blood draws without substituting CGM glucose. Fasting insulin analyses used draws from March 2023 onward. BMI was computed from height and the closest logged weight within ±90 days (restricting to 15–60 kg/m²).

The primary laboratory cohort (n = 7,930) excluded any participant with a diabetes-range laboratory value (HbA1c ≥6.5% or fasting glucose ≥126 mg/dL) at any point in their record. All analyses were complete-case without imputation (sample sizes in Table 1). Reliability and meal analyses included participants without laboratory data (excluding any participant with a known diabetes-range lab value; participants without laboratory results could not be screened).

### Statistical analysis

Week-to-week CGM reliability was evaluated across each participant’s first two non-overlapping 7-day blocks of consecutive dense days using the two-way, absolute-agreement intraclass correlation, ICC(A,1);^35^ wear-duration reliability compared two k-day windows separated by 14 days. Same-food repeatability was the Pearson correlation (r) across person-food pairs between the mean 2-hour iAUCs of the first k and next k isolated meals in chronological order (2,000 participant-level bootstrap resamples). Carbohydrate-sensitivity repeatability was the correlation between slopes fitted separately to each participant’s odd– and even-numbered meals (≥30 meals per half), while comparisons with insulin resistance used slopes fitted to meals within ±90 days of the anchor draw.

Laboratory markers (fasting insulin, HOMA-IR, triglycerides, triglyceride-to-HDL ratio, HDL cholesterol, uric acid, hsCRP, ApoB, and LDL cholesterol) were log-transformed and regressed on standardized CGM metrics via ordinary least squares with heteroskedasticity-consistent (HC3) 95% CIs, reported as percentage differences per SD. Markers were log-transformed because fasting insulin, HOMA-IR, triglycerides, and hsCRP are strongly right-skewed; transforming every marker puts all associations on the same percentage scale. Models controlling for average glucose included standardized average glucose as the sole covariate. Adjustment for age decade and sex altered primary estimates by at most 0.84 percentage points (Supplementary Table 7), and BMI was evaluated in BMI-complete participants. Single-predictor variance explained (R²) by average glucose versus HbA1c was compared using 2,000 paired participant-bootstrap resamples overall, within HbA1c <5.7%, and across the other laboratory markers, for which percentage differences per SD were adjusted for age decade and sex (Supplementary Table 1a). Within HbA1c <5.7%, we compared the prevalence of elevated fasting insulin (≥10 µIU/mL, above the cohort upper quartile; Table 1) above versus below an average glucose of 110 mg/dL (above normative reference cohorts;^13^ both on the CGM sensor scale), estimated the area under the curve for that insulin threshold, and quantified incremental R² when adding each glycemic measure to the other.

Robustness analyses repeated the HbA1c comparisons by anchor year and within average glucose <110 mg/dL (Supplementary Table 3); quantified laboratory test-retest reliability (ICC(1)) across repeat blood draws 14 to 90 days apart (Supplementary Table 4); adjusted spike-width models for BMI and, separately, for spike height and frequency (Fig. 4d); evaluated HbA1c across age decades at fixed average glucose adjusted for sex and anchor year (Supplementary Fig. 3); and compared cohort HbA1c-glucose regressions against the published GMI slope^17^ (Supplementary Fig. 1). Between-group differences (Fig. 5e; Supplementary Table 1b) were ratios of geometric means from regression of the log marker on group. False-discovery rates were controlled within marker families; all tests were two-sided, with bootstrap contrast P-values derived via normal approximation.

### Use of artificial intelligence

AI models assisted with code, number checking, and editing under author direction. The authors reviewed all code and text and take responsibility for the content.

## Data availability

Participant-level data are not publicly shareable under the consent terms. The aggregate source data underlying each figure and table are available in the code repository (see Code availability). The authors are open to research collaborations; requests can be directed to the corresponding author.

## Code availability

Analysis and figure code, and the source data for each figure and table, are available at https://github.com/PrecisionLongevity/levels-cgm-paper-code.

## Supporting information

STROBE Checklist

## Acknowledgements

We thank Robert Lustig and Josh Clemente for comments and feedback on the manuscript, and the participants who shared their data.

## Author contributions

P.G. and M.L. contributed equally to conceptualization, formal analysis, and writing (original draft, review and editing). Both authors read and approved the manuscript.

## Supplementary Information

- **Supplementary Tables:**
  ◦ 1: average glucose and HbA1c against other laboratory markers of metabolic health (a) and in the HbA1c and average glucose groups (b);
  ◦ 2: spike width in responses to logged meals;
  ◦ 3: HbA1c comparison by anchor year and with the sample restricted on average glucose;
  ◦ 4: test-retest reliability of laboratory measures;
  ◦ 5: metric definitions;
  ◦ 6: laboratory units, conversions, and accepted ranges;
  ◦ 7: age and sex sensitivity.
- **Supplementary Figs.:**
  ◦ 1: HbA1c and average glucose: the cohort reproduces the published GMI slope;
  ◦ 2: HbA1c–average glucose discordant groups differ in body weight, diet, and lifestyle;
  ◦ 3: HbA1c by age at the same average glucose.

