## Supplementary Info for "Consumer continuous glucose monitoring uncovers hidden metabolic risk in free-living adults without diabetes"

#### Contents

Supplementary Tables 1–7

Supplementary Figs. 1–3

### Supplementary Tables

**Supplementary Table 1a | Average glucose and HbA1c against other laboratory markers of metabolic health.** Average glucose explained more of the variance than HbA1c for seven of the nine markers and stayed associated with all seven after adjustment for BMI; for ApoB and LDL cholesterol, both measures showed small associations.

| Marker | Participants | $R^2$ average glucose | $R^2$ HbA1c | Difference (95% CI) | Average glucose, % per SD (95% CI) | HbA1c, % per SD (95% CI) | Average glucose + BMI, % per SD (95% CI) | Participants with BMI |
|---|---|---|---|---|---|---|---|---|
| Fasting insulin | 6,890 | 0.131 | 0.048 | 0.083 (0.068 to 0.097) | 29.2 (27.1 to 31.3) | 17.7 (15.6 to 19.8) | 16.6 (14.3 to 18.9) | 4,252 |
| HOMA-IR | 1,192 | 0.143 | 0.059 | 0.084 (0.046 to 0.124) | 30.1 (25.3 to 35.0) | 19.9 (15.1 to 24.8) | 16.4 (10.5 to 22.6) | 713 |
| Triglycerides | 7,804 | 0.106 | 0.029 | 0.077 (0.064 to 0.090) | 14.7 (13.6 to 15.9) | 7.2 (6.1 to 8.3) | 9.3 (7.9 to 10.7) | 4,724 |
| HDL cholesterol | 2,139 | 0.067 | 0.009 | 0.058 (0.039 to 0.079) | -6.0 (-7.0 to -4.9) | -3.0 (-4.1 to -1.8) | -1.8 (-3.2 to -0.3) | 1,208 |
| Triglyceride-to-HDL ratio | 2,137 | 0.099 | 0.025 | 0.074 (0.051 to 0.099) | 20.9 (17.6 to 24.3) | 10.9 (7.9 to 13.9) | 9.0 (5.1 to 13.0) | 1,207 |
| Uric acid | 6,503 | 0.086 | 0.020 | 0.066 (0.055 to 0.079) | 5.6 (5.0 to 6.2) | 3.0 (2.4 to 3.7) | 3.5 (2.7 to 4.3) | 4,030 |
| hsCRP | 1,171 | 0.030 | 0.007 | 0.023 (0.005 to 0.045) | 28.5 (19.7 to 37.9) | 9.2 (0.9 to 18.2) | 10.9 (0.4 to 22.4) | 650 |
| ApoB | 6,741 | 0.006 | 0.015 | -0.009 (-0.015 to -0.004) | 1.8 (1.1 to 2.5) | 2.6 (1.8 to 3.3) | 0.6 (-0.3 to 1.4) | 4,176 |
| LDL cholesterol | 2,152 | 0.002 | 0.004 | -0.002 (-0.009 to 0.004) | 2.0 (0.3 to 3.8) | 2.7 (1.0 to 4.5) | 2.1 (-0.2 to 4.5) | 1,212 |

Primary cohort (no diabetes-range laboratory value). Each marker is the canonical panel value nearest the anchor HbA1c draw (within 7 days), log-transformed. $R^2$: single-predictor variance explained in the same participants; difference: paired participant bootstrap 95% CI (2,000 resamples). % per SD: OLS adjusted for age decade and sex (and BMI where stated), HC3 95% CIs.

**Supplementary Table 1b | Laboratory markers in the HbA1c and average glucose groups.** Participants with elevated average glucose and a normal HbA1c had higher fasting insulin (19.9%), HOMA-IR (18.9%), and triglycerides (18.2%) than those with an elevated HbA1c and normal average glucose; hsCRP was 27.2% higher, with a 95% CI that included 0.

| Marker | Both normal, GM (n) | CGM elevated only, GM (n) | HbA1c elevated only, GM (n) | Both elevated, GM (n) | CGM-only vs HbA1c-only, % (95% CI) |
|---|---|---|---|---|---|
| Fasting insulin | 4.92 (3,692) | 6.86 (1,087) | 5.72 (1,100) | 8.96 (1,011) | 19.9 (13.3 to 26.8) |
| HOMA-IR | 1.18 (683) | 1.66 (201) | 1.40 (162) | 2.24 (146) | 18.9 (3.0 to 37.2) |
| Triglycerides | 70.06 (4,140) | 87.87 (1,280) | 74.34 (1,251) | 97.46 (1,133) | 18.2 (14.2 to 22.4) |

| Marker | Both normal, GM (n) | CGM elevated only, GM (n) | HbA1c elevated only, GM (n) | Both elevated, GM (n) | CGM-only vs HbA1c-only, % (95% CI) |
|---|---|---|---|---|---|
| hsCRP | 0.87 (686) | 1.10 (228) | 0.86 (140) | 1.53 (117) | 27.2 (-0.8 to 63.2) |
| Fasting insulin >=10 µIU/mL, % | 12.6 (11.5 to 13.7) | 27.5 (24.9 to 30.2) | 19.6 (17.4 to 22.1) | 44.5 (41.5 to 47.6) | |
| HOMA-IR >=2.5, % | 11.3 (9.1 to 13.9) | 24.4 (19.0 to 30.8) | 16.0 (11.2 to 22.5) | 44.5 (36.7 to 52.6) | |

CGM elevated: average glucose >=110 mg/dL; HbA1c elevated: >=5.7% (rounded half-up to 0.1). GM, geometric mean (µIU/mL, HOMA-IR units, mg/dL, mg/L). Ratio: OLS of the log marker on group, HC3, unadjusted. Threshold rows: share above the threshold, Wilson 95% CI.

**Supplementary Table 2 | Spike width replicated in responses to logged meals, in participants with both widths.** In the same 3,484 participants, the width of responses to logged meals was associated with fasting insulin as strongly as CGM spike width (8.0% vs 8.1% per SD) and remained associated after adjustment for meal content (7.3%), so the width signal does not depend on how spikes are detected.

| Width measure | Compared at the | Participants | Fasting insulin, % per SD (95% CI) | Participants with HOMA-IR | HOMA-IR, % per SD (95% CI) |
|---|---|---|---|---|---|
| CGM spike width | same average glucose | 3,484 | 8.1 (5.8 to 10.5) | 841 | 5.4 (0.6 to 10.4) |
| Width of responses to logged meals | same average glucose | 3,484 | 8.0 (5.8 to 10.3) | 841 | 5.5 (0.7 to 10.5) |
| Width of responses to logged meals, adjusted for meal content and context | same average glucose | 3,484 | 7.3 (5.0 to 9.7) | 841 | 4.6 (-0.4 to 9.8) |
| CGM spike width | same average glucose, both widths in one model | 3,484 | 5.1 (2.4 to 7.9) | 841 | 3.3 (-2.1 to 9.0) |
| Width of responses to logged meals | same average glucose, both widths in one model | 3,484 | 4.8 (2.2 to 7.6) | 841 | 3.5 (-2.0 to 9.3) |

Participants with both a CGM spike width and a width of responses to logged meals (fasting insulin, 3,484; HOMA-IR, 841). Width of responses to logged meals: isolated logged meals with a 2-h rise >= 20 mg/dL, minutes above the pre-meal baseline + half the rise, participant mean over >= 10 meals; adjusted: participant mean of meal-level residuals after regressing width on macronutrients (linear and squared), log peak rise, pre-meal glucose, meal-hour band, fasting for 4 h, and exercise within 2 h. Log outcome per SD with average glucose as the covariate; in the one-model rows both widths and average glucose are in the same model. HC3 95% CIs.

**Supplementary Table 3 | HbA1c comparison by anchor year and with the sample restricted on average glucose.** Average glucose explained more of the variance in fasting insulin than HbA1c in every anchor year and when the sample was restricted on average glucose instead of HbA1c, and discriminated fasting insulin of 10 µIU/mL or higher better in every year (95% CIs of the differences above 0).

| Sample | Participants | $R^2$ average glucose | $R^2$ HbA1c | Difference (95% CI) | AUC average glucose | AUC HbA1c | AUC difference (95% CI) |
|---|---|---|---|---|---|---|---|
| Anchor year 2023 | 1442 | 0.111 | 0.052 | 0.059 (0.026 to 0.090) | 0.71 | 0.64 | 0.07 (0.03 to 0.11) |
| Anchor year 2024 | 3789 | 0.135 | 0.048 | 0.087 (0.065 to 0.107) | 0.70 | 0.64 | 0.05 (0.03 to 0.08) |

| Sample | Participants | $R^2$ average glucose | $R^2$ HbA1c | Difference (95% CI) | AUC average glucose | AUC HbA1c | AUC difference (95% CI) |
|---|---|---|---|---|---|---|---|
| Anchor year 2025 | 1657 | 0.135 | 0.041 | 0.093 (0.063 to 0.124) | 0.69 | 0.61 | 0.08 (0.04 to 0.11) |
| Average glucose <110 mg/dL | 4792 | 0.055 | 0.012 | 0.043 (0.031 to 0.055) | | | |

$R^2$: single-predictor variance in log fasting insulin; AUC: fasting insulin >=10 μIU/mL. Paired bootstrap 95% CIs (2,000 resamples). Two participants anchored outside 2023-2025 are not shown.

**Supplementary Table 4 | Test-retest reliability of laboratory measures.** HbA1c and fasting insulin were reproducible in draws 14 to 90 days apart (ICC 0.72 and 0.71), so the weak resolution of HbA1c in the normal range does not reflect assay noise; a single laboratory fasting glucose was less reproducible (0.51).

| Measure | Pairs | Median days between draws | ICC(1) | Pearson r |
|---|---|---|---|---|
| HbA1c | 607 | 63 | 0.72 | 0.74 |
| Fasting insulin (log) | 449 | 65 | 0.71 | 0.72 |
| HOMA-IR (log) | 71 | 64 | 0.53 | 0.55 |
| Laboratory fasting glucose | 194 | 58 | 0.51 | 0.51 |

First pair of draws 14 to 90 days apart per participant, among participants never in the diabetes range. One-way ICC. CGM average glucose week-to-week ICC(A,1) was 0.747 for comparison.

**Supplementary Table 5 | Continuous glucose measures: definitions, units, and distributions.**

| Measure | Definition | Unit | Cohort median (IQR) | Summarized over |
|---|---|---|---|---|
| Average glucose | average of the observed 15-minute cells of the record | mg/dL | 106.7 (100.7 to 113.4) | whole record; 49,876 participants |
| SD | standard deviation of the same cells (spread around the average) | mg/dL | 16.6 (14.4 to 19.2) | whole record; 49,692 participants |
| CV | SD divided by the average, × 100 | % | 15.6 (13.8 to 17.6) | whole record; 49,692 participants |
| MODD | mean absolute difference between consecutive days at the same clock time | mg/dL | 12.9 (11.2 to 14.9) | lab-linked panel; 7,930 participants |
| MAGE | mean amplitude of glycemic excursions: mean of rises and falls larger than 1 SD (moving-average algorithm; Methods) | mg/dL | 35.9 (31.2 to 41.7) | lab-linked panel; 7,930 participants |
| Time 70–140 mg/dL | share of observed time with glucose between 70 and 140 mg/dL | % of time | 94.6 (90.8 to 96.8) | whole record; 49,876 participants |
| Spike frequency | spikes per eligible day; a spike is a peak ≥30 mg/dL above the lowest value of the preceding 90 min | spikes per day | 4.0 (2.9 to 5.2) | lab-linked panel; 7,914 participants |

| Measure | Definition | Unit | Cohort median (IQR) | Summarized over |
|---|---|---|---|---|
| Spike height | a participant's mean rise across their spikes: peak minus its preceding 90-min low | mg/dL | 43.3 (41.0 to 46.0) | lab-linked panel; 7,914 participants |
| Spike width | minutes above half the rise of each spike, participant mean over resolved spikes | min | 65.9 (60.1 to 73.1) | lab-linked panel; 7,907 participants |
| Meal peak rise | highest value 0–120 min after a logged meal minus the pre-meal baseline | mg/dL | 22 (12 to 36) | isolated logged meals; 751,450 meals |
| 2-h iAUC | area between the curve and the baseline, 0–120 min, counting only area above it | mg/dL·min | 980 (380 to 1,895) | isolated logged meals; 751,450 meals |
| Carbohydrate sensitivity | a person's peak rise per gram of carbohydrate, adjusted for fiber, fat, protein | mg/dL per g | 0.33 (0.20 to 0.49) | ≥30 logged meals; 11,332 participants |

Whole record = every participant's CGM record (observed 15-minute cells, accepted readings 20–500 mg/dL). Lab-linked panel = ±90 days around the canonical HbA1c draw, primary cohort without an observed diabetes-range laboratory value. Isolated logged meals and carbohydrate slopes exclude the 412 participants with any diabetes-range laboratory value. MODD keeps its conventional name (mean of daily differences). IQR, interquartile range. Spikes are detected from the CGM trace alone; meal peak rise and 2-h iAUC are measured only after isolated logged meals.

**Supplementary Table 6 | Laboratory units, conversions, and accepted ranges.**

| Analyte | Unit | Accepted range | Accepted alternate units and conversion |
|---|---|---|---|
| HbA1c | % | 3–20 | IFCC mmol/mol ×0.09148 +2.152 |
| Fasting glucose | mg/dL | 30–600 | mmol/L ×18.0182 |
| Fasting insulin | μIU/mL | 0.5–500 | μU/mL, u[IU]/mL, mcIU/mL, and mU/L are equivalent |
| Triglycerides | mg/dL | 10–5,000 | mmol/L ×88.57 |
| HDL cholesterol | mg/dL | 5–200 | mmol/L ×38.67 |
| LDL cholesterol | mg/dL | 10–500 | mmol/L ×38.67 |
| ApoB | mg/dL | 20–250 | g/L ×100 |
| hsCRP | mg/L | 0.01–100 | mg/dL ×10 |
| Uric acid | mg/dL | 1–20 | None |

Units were normalized for case, spacing, and the micro sign and checked against this list; a unit was never inferred from the size of a value. Conversions were applied before the range check. HbA1c reported as % of total hemoglobin and cholesterol reported as mg/dL (calculated) were also accepted. Insulin reported in pmol/L was excluded because no assay-specific conversion was available. The ranges are limits of analytic validity, not clinical reference intervals; values outside them were excluded before same-day values were combined. ApoB, apolipoprotein B; HDL, high-density lipoprotein; hsCRP, high-sensitivity C-reactive protein; IFCC, International Federation of Clinical Chemistry; LDL, low-density lipoprotein.

**Supplementary Table 7 | Age and sex sensitivity.** Adjustment for age and sex changed no estimate by more than 0.84 percentage points, and beyond age and sex, average glucose still explained more of the variance in fasting insulin than HbA1c (13.2% vs 4.9%).

#### A | Per-SD associations

| Comparison | Marker | Measure | n | % per SD, without age/sex (95% CI) | % per SD, with age/sex (95% CI) | change (points) |
|---|---|---|---|---|---|---|
| unadjusted → + age, sex | HOMA-IR | Average glucose | 1,192 | +30.0 (+25.3 to +34.9) | +30.3 (+25.5 to +35.2) | +0.26 |
| unadjusted → + age, sex | HOMA-IR | SD | 1,192 | +10.8 (+6.3 to +15.5) | +10.6 (+6.1 to +15.3) | −0.17 |
| unadjusted → + age, sex | HOMA-IR | CV | 1,192 | +0.4 (−3.9 to +5.0) | +0.3 (−4.1 to +4.9) | −0.14 |
| unadjusted → + age, sex | HOMA-IR | MODD | 1,192 | +13.7 (+9.4 to +18.1) | +13.7 (+9.4 to +18.2) | +0.01 |
| unadjusted → + age, sex | HOMA-IR | MAGE | 1,192 | +7.9 (+3.5 to +12.6) | +7.8 (+3.3 to +12.5) | −0.17 |
| unadjusted → + age, sex | HOMA-IR | Spike frequency | 1,189 | +7.8 (+3.5 to +12.2) | +8.3 (+3.9 to +12.9) | +0.57 |
| unadjusted → + age, sex | HOMA-IR | Spike height | 1,189 | +5.4 (+0.4 to +10.6) | +5.2 (+0.2 to +10.5) | −0.15 |
| unadjusted → + age, sex | Fasting insulin | Average glucose | 6,890 | +28.3 (+26.2 to +30.4) | +29.1 (+27.0 to +31.3) | +0.84 |
| unadjusted → + age, sex | Fasting insulin | SD | 6,890 | +10.1 (+8.2 to +12.1) | +10.0 (+8.1 to +12.0) | −0.08 |
| unadjusted → + age, sex | Fasting insulin | CV | 6,890 | −0.6 (−2.3 to +1.2) | −0.5 (−2.3 to +1.2) | +0.01 |
| unadjusted → + age, sex | Fasting insulin | MODD | 6,890 | +13.4 (+11.4 to +15.5) | +13.4 (+11.4 to +15.5) | +0.01 |
| unadjusted → + age, sex | Fasting insulin | MAGE | 6,890 | +8.9 (+7.0 to +10.9) | +8.8 (+6.9 to +10.8) | −0.11 |
| unadjusted → + age, sex | Fasting insulin | Spike frequency | 6,882 | +7.4 (+5.6 to +9.3) | +8.2 (+6.3 to +10.1) | +0.78 |
| unadjusted → + age, sex | Fasting insulin | Spike height | 6,882 | +7.0 (+5.1 to +8.9) | +7.0 (+5.1 to +9.0) | +0.03 |
| same average glucose → + age, sex | HOMA-IR | SD | 1,192 | −3.0 (−7.4 to +1.6) | −3.1 (−7.5 to +1.5) | −0.12 |
| same average glucose → + age, sex | HOMA-IR | CV | 1,192 | −2.7 (−6.6 to +1.4) | −2.8 (−6.8 to +1.3) | −0.12 |
| same average glucose → + age, sex | HOMA-IR | MODD | 1,192 | −1.9 (−6.0 to +2.4) | −1.9 (−6.1 to +2.4) | −0.03 |
| same average glucose → + age, sex | HOMA-IR | MAGE | 1,192 | −6.5 (−10.1 to −2.7) | −6.7 (−10.3 to −2.9) | −0.21 |
| same average glucose → + age, sex | HOMA-IR | Spike frequency | 1,189 | −5.4 (−9.4 to −1.2) | −5.6 (−9.8 to −1.2) | −0.13 |
| same average glucose → + age, sex | HOMA-IR | Spike height | 1,189 | −6.2 (−9.7 to −2.7) | −6.8 (−10.2 to −3.2) | −0.54 |
| same average glucose → + age, sex | Fasting insulin | SD | 6,890 | −4.8 (−6.6 to −3.0) | −4.9 (−6.7 to −3.0) | −0.07 |
| same average glucose → + age, sex | Fasting insulin | CV | 6,890 | −4.3 (−5.8 to −2.7) | −4.3 (−5.9 to −2.8) | −0.05 |
| same average glucose → + age, sex | Fasting insulin | MODD | 6,890 | −2.3 (−4.2 to −0.3) | −2.4 (−4.3 to −0.4) | −0.14 |

| Comparison | Marker | Measure | n | % per SD, without age/sex (95% CI) | % per SD, with age/sex (95% CI) | change (points) |
|---|---|---|---|---|---|---|
| same average glucose → + age, sex | Fasting insulin | MAGE | 6,890 | −6.3 (−8.0 to −4.5) | −6.6 (−8.3 to −4.8) | −0.28 |
| same average glucose → + age, sex | Fasting insulin | Spike frequency | 6,882 | −4.8 (−6.5 to −3.1) | −5.1 (−6.8 to −3.4) | −0.29 |
| same average glucose → + age, sex | Fasting insulin | Spike height | 6,882 | −5.1 (−6.7 to −3.4) | −5.6 (−7.2 to −3.9) | −0.50 |
| unadjusted → + age, sex | HOMA-IR | Pre-meal glucose | 814 | +21.1 (+15.3 to +27.1) | +21.4 (+15.4 to +27.7) | +0.37 |
| unadjusted → + age, sex | HOMA-IR | Meal peak rise | 814 | +3.6 (−1.0 to +8.4) | +3.4 (−1.3 to +8.3) | −0.17 |
| unadjusted → + age, sex | HOMA-IR | Meal 2-h iAUC | 814 | +5.5 (+0.6 to +10.6) | +5.1 (+0.2 to +10.3) | −0.40 |
| unadjusted → + age, sex | Fasting insulin | Pre-meal glucose | 3,503 | +18.6 (+15.9 to +21.2) | +19.1 (+16.4 to +21.9) | +0.56 |
| unadjusted → + age, sex | Fasting insulin | Meal peak rise | 3,503 | +3.4 (+1.1 to +5.7) | +3.5 (+1.2 to +5.9) | +0.17 |
| unadjusted → + age, sex | Fasting insulin | Meal 2-h iAUC | 3,503 | +5.3 (+2.9 to +7.7) | +5.3 (+2.9 to +7.7) | −0.01 |
| same average glucose → + age, sex | HOMA-IR | Pre-meal glucose | 814 | −10.6 (−18.2 to −2.3) | −10.5 (−18.2 to −2.1) | +0.10 |
| same average glucose → + age, sex | HOMA-IR | Meal peak rise | 814 | −5.7 (−9.9 to −1.2) | −6.0 (−10.2 to −1.5) | −0.30 |
| same average glucose → + age, sex | HOMA-IR | Meal 2-h iAUC | 814 | −3.1 (−7.5 to +1.5) | −3.6 (−8.0 to +1.0) | −0.49 |
| same average glucose → + age, sex | Fasting insulin | Pre-meal glucose | 3,503 | −11.3 (−14.8 to −7.7) | −11.4 (−15.0 to −7.7) | −0.09 |
| same average glucose → + age, sex | Fasting insulin | Meal peak rise | 3,503 | −5.1 (−7.2 to −3.0) | −5.2 (−7.3 to −3.0) | −0.09 |
| same average glucose → + age, sex | Fasting insulin | Meal 2-h iAUC | 3,503 | −2.8 (−4.9 to −0.6) | −3.1 (−5.2 to −0.9) | −0.29 |

#### B | Variance explained

| Outcome | Scope | n | $R^2$ average glucose, unadjusted (%) | $R^2$ HbA1c, unadjusted (%) | $\Delta R^2$ average glucose beyond age/sex (%) | $\Delta R^2$ HbA1c beyond age/sex (%) | difference (points) |
|---|---|---|---|---|---|---|---|
| Fasting insulin | whole primary cohort | 6,890 | 13.1 | 4.8 | 13.2 (11.7–14.7) | 4.9 (3.9–6.0) | +8.3 (+6.8 to +9.8) |
| Fasting insulin | HbA1c <5.7% | 4,779 | 8.9 | 0.5 | 9.1 (7.5–10.7) | 0.6 (0.2–1.1) | +8.5 (+6.9 to +10.0) |
| HOMA-IR | whole primary cohort | 1,192 | 14.3 | 5.9 | 14.2 (10.7–18.0) | 6.0 (3.7–8.9) | +8.2 (+4.0 to +12.2) |
| HOMA-IR | HbA1c <5.7% | 884 | 11.0 | 2.2 | 10.6 (7.1–14.5) | 2.1 (0.7–4.1) | +8.5 (+4.6 to +12.5) |

Primary estimates in the paper are unadjusted; this table refits them with age bucket and sex. A, Per-SD % difference in HOMA-IR and fasting insulin, same participants without and with age and sex (maximum absolute change 0.84 points). B, Variance in log fasting insulin and log HOMA-IR explained by average glucose and by HbA1c, unadjusted and beyond age and sex (participant-bootstrap 95% CI, 2,000 resamples).

### Supplementary Figures

**Supplementary Fig. 1 | HbA1c and average glucose: the cohort reproduces the published GMI slope.**

**Supplementary Fig. 1 | HbA1c and average glucose: the cohort reproduces the published GMI slope**

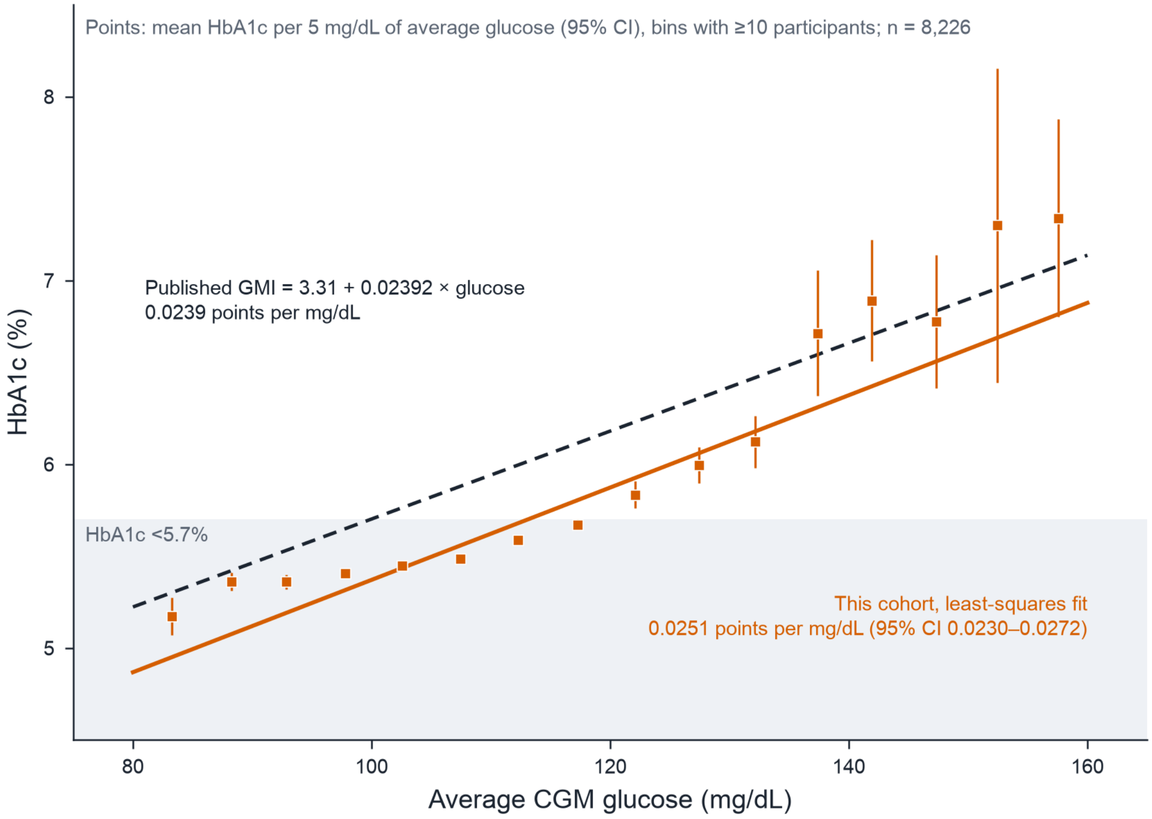


HbA1c rose with average glucose at the published GMI slope (0.0251 percentage points per mg/dL in this cohort, 95% CI 0.0230 to 0.0272, against 0.0239), so these consumer CGM data relate to HbA1c as clinical CGM data do. Mean HbA1c in each 5 mg/dL bin of average CGM glucose (bins with at least 10 participants) in all 8,226 participants with paired HbA1c and CGM data, including 296 with a diabetes-range laboratory value. Solid line, least-squares fit of HbA1c on average glucose in this cohort (0.0251 percentage points per mg/dL); dashed line, the published glucose management indicator (GMI = 3.31 + 0.02392 × average glucose; ref. 17). Shading, HbA1c below 5.7%. Error bars, 95% confidence intervals. CGM, continuous glucose monitoring; GMI, glucose management indicator.

**Supplementary Fig. 2 | HbA1c–average glucose discordant groups differ in body weight, diet, and lifestyle.**

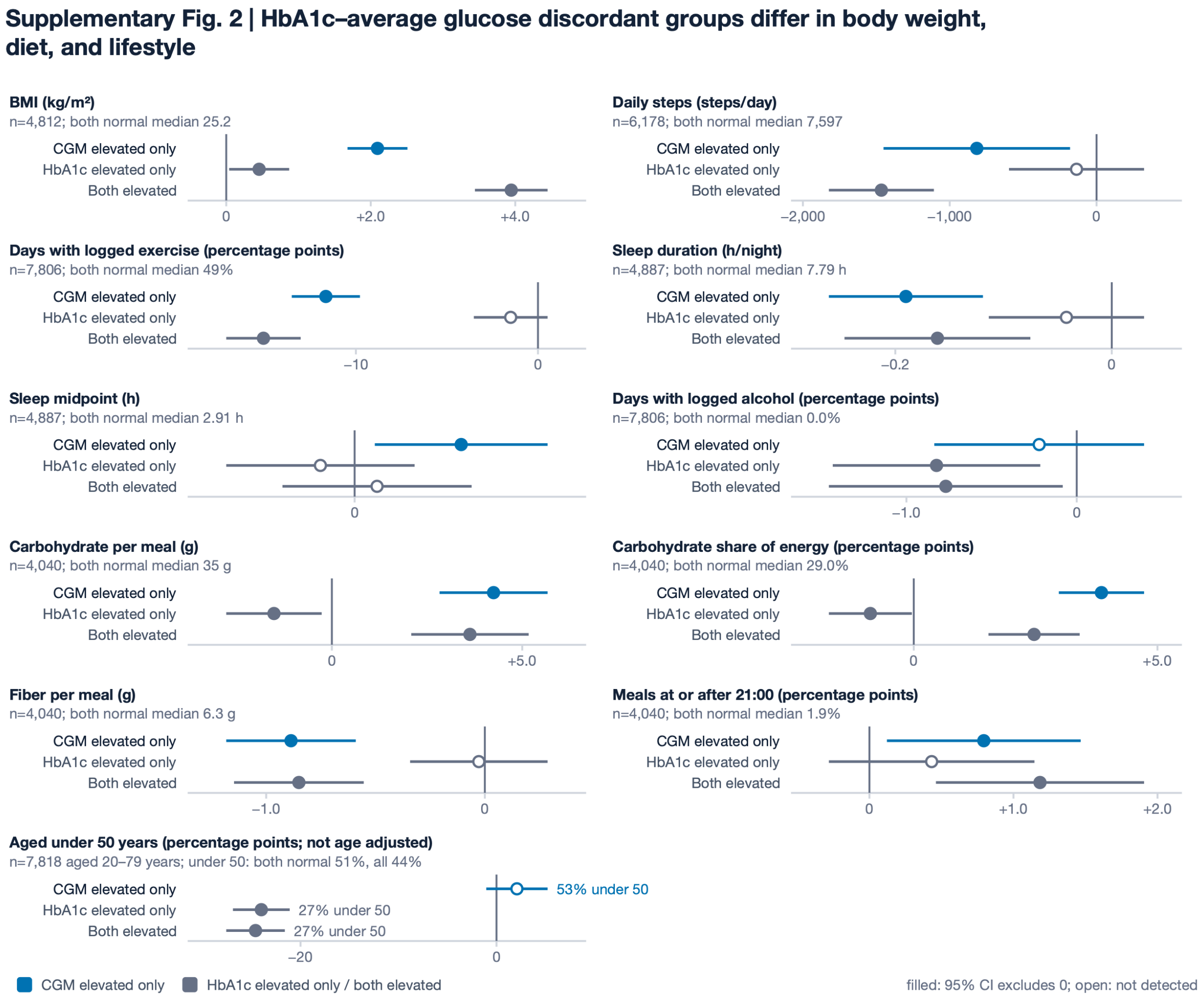


Participants with elevated average glucose and a normal HbA1c were younger than those with an elevated HbA1c only (53% vs 27% under 50 years) and, adjusted for age and sex, were heavier, less active, slept less, and ate more carbohydrate and less fiber per meal than participants normal on both measures. Difference from participants normal on both measures for the groups with elevated average glucose only (≥110 mg/dL), elevated HbA1c only (≥5.7%), and both elevated, in the primary cohort (7,930 participants). Above each panel, the number of participants with the measure and the median in the both-normal group. The first ten panels are adjusted for age and sex; the last shows the share aged under 50 years. Activity and sleep are from CGM days within ±90 days of the HbA1c draw (at least 7 days); diet is from at least 10 isolated logged meals. Filled markers, 95% confidence interval excludes 0; open markers, interval includes 0. Error bars, 95% confidence intervals. BMI, body mass index; CGM, continuous glucose monitoring.

**Supplementary Fig. 3 | HbA1c by age at the same average glucose.**

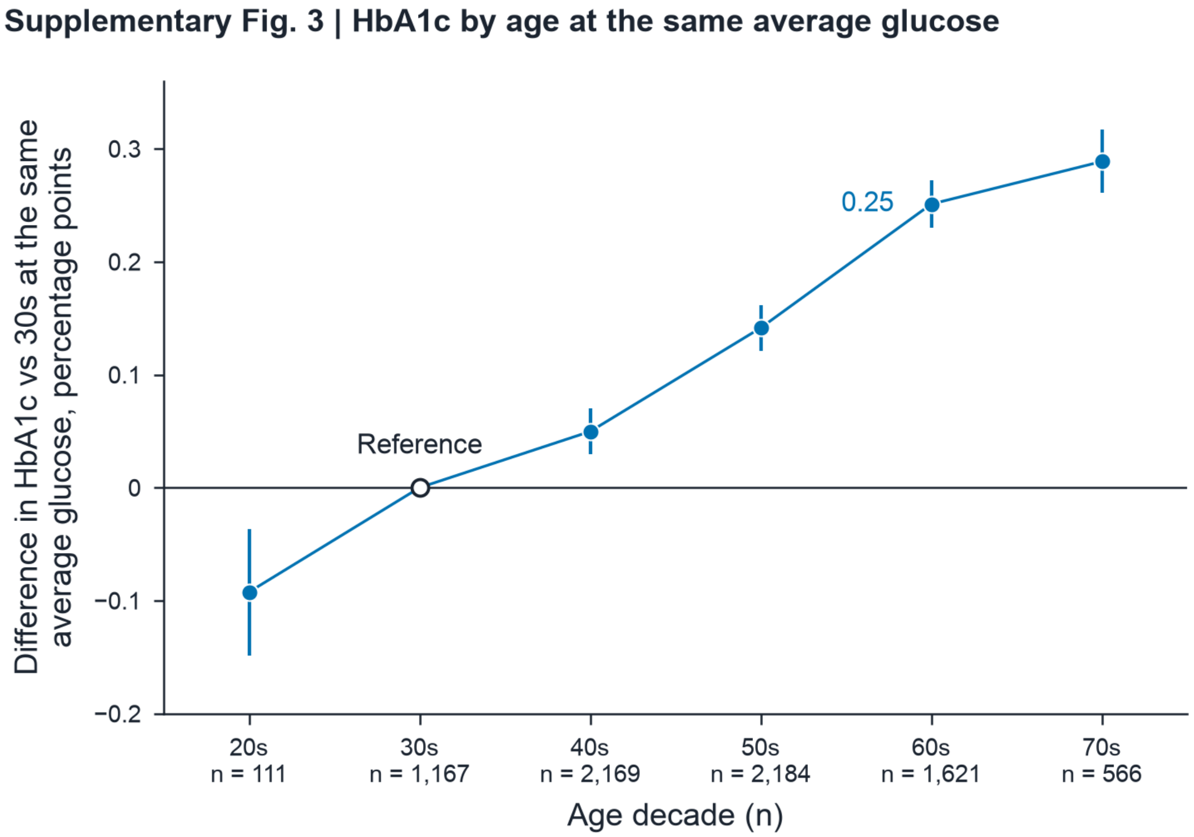


At the same average glucose, HbA1c was higher in older participants (0.25 percentage points higher in their 60s than in their 30s), so a fixed HbA1c cutoff of 5.7% is reached at a lower average glucose in older than in younger adults. Difference in HbA1c between participants in each age decade and those in their 30s (open marker, reference), from linear regression of HbA1c on age decade with average CGM glucose, sex, and anchor year as covariates, in 7,818 primary-cohort participants aged 20–79 years (number per decade below the axis). The labelled value is for participants in their 60s. Error bars, 95% confidence intervals. CGM, continuous glucose monitoring.
