## Supplementary material for "Consumer continuous glucose monitoring uncovers hidden metabolic risk in free-living adults without diabetes": STROBE Checklist

**STROBE checklist (cross-sectional studies)**

Ginart & Loesche. Consumer continuous glucose monitoring uncovers hidden metabolic risk in free-living adults without diabetes. Manuscript version 11.9. Checklist items follow the STROBE statement (von Elm et al., Lancet 2007; reference 31 of the manuscript). *Items 8 and 13-15 apply to the single cross-sectional sample. Locations refer to manuscript sections, figures and tables.

| Section/topic | Item | Recommendation (summary) | Where reported |
|---|---|---|---|
| Title and abstract | 1 | (a) Indicate the study design with a commonly used term in the title or abstract; (b) provide an informative, balanced summary of what was done and found | Abstract ("In this cross-sectional study of 62,122 free-living adults...") |
| Background/rationale | 2 | Explain the scientific background and rationale | Introduction, paragraphs 1-3 |
| Objectives | 3 | State specific objectives, including prespecified hypotheses | Introduction, paragraphs 2-3 (wear duration, measures that track insulin resistance, risk missed by a normal HbA1c, post-meal features) |
| Study design | 4 | Present key elements of study design early in the paper | Methods: Study design and participants (cross-sectional analysis; protocol LH-001) |
| Setting | 5 | Describe the setting, locations and relevant dates, including recruitment, exposure, follow-up and data collection | Methods: Study design and participants; Platform and data streams (US direct-to-consumer platform, 2022-2025) |
| Participants | 6 | Give eligibility criteria and the sources and methods of selection of participants | Methods: Study design and participants (eligibility, exclusions); Laboratory pairing (primary laboratory cohort); Fig. 1b |
| Variables | 7 | Clearly define all outcomes, exposures, predictors, potential confounders and effect modifiers | Methods: CGM measures; Logged meals; Laboratory pairing; Statistical analysis; Supplementary Table 5 (measure definitions) |
| Data sources/ measurement | 8* | For each variable, give sources of data and details of methods of assessment | Methods: Platform and data streams; CGM measures; Laboratory pairing; Supplementary Table 6 (laboratory units and accepted ranges) |
| Bias | 9 | Describe any efforts to address potential sources of bias | Methods: Laboratory pairing (diabetes-range exclusion), Statistical analysis (robustness analyses); Discussion: limitations paragraph |
| Study size | 10 | Explain how the study size was arrived at | All consenting participants meeting each analysis criterion were included; no sample size was prespecified (Results: first section; Fig. 1b) |
| Quantitative variables | 11 | Explain how quantitative variables were handled; if applicable, describe which groupings were chosen and why | Methods: Statistical analysis (per-SD standardization, log-transformed markers, thresholds 110 mg/dL, HbA1c 5.7% and fasting insulin 10 μIU/mL with rationale) |
| Statistical methods | 12 | (a) Describe all statistical methods, including those used to control for confounding; (b) methods for subgroups and interactions; (c) how missing data were addressed; (d) analytic methods taking account of sampling strategy, if applicable; (e) sensitivity analyses | (a) Methods: Statistical analysis (OLS with HC3 CIs; adjustment for age decade, sex and BMI); (b) within HbA1c <5.7% and discordant-group analyses; (c) complete-case analysis (Methods: Laboratory pairing); (d) not applicable (no sampling design); (e) Methods: Statistical analysis, robustness paragraph; Supplementary Tables 3, 4 and 7 |

| Section/topic | Item | Recommendation (summary) | Where reported |
|---|---|---|---|
| Participants | 13* | (a) Report numbers at each stage of study; (b) give reasons for non-participation at each stage; (c) consider use of a flow diagram | (a) Results: first section; Fig. 1b; (b) Fig. 1b (data streams contributed; diabetes-range exclusion); (c) Fig. 1b (flow from consent to the primary cohort) |
| Descriptive data | 14* | (a) Give characteristics of study participants; (b) indicate the number of participants with missing data for each variable of interest | (a) Table 1; Fig. 1c-e; (b) Table 1 (n per variable) |
| Outcome data | 15* | Report numbers of outcome events or summary measures | Results; Table 1; Figs 3-6 |
| Main results | 16 | (a) Give unadjusted estimates and, if applicable, confounder-adjusted estimates and their precision (e.g. 95% CI); (b) report category boundaries when continuous variables were categorized; (c) consider translating relative risk into absolute risk | (a) Results; Figs 3-5 (unadjusted and age/sex- and BMI-adjusted estimates with 95% CIs); Supplementary Tables 1a and 7; (b) Fig. 3a (quartile boundaries), Fig. 5d (thresholds); (c) prevalences in Fig. 3a and Fig. 5d |
| Other analyses | 17 | Report other analyses done, e.g. analyses of subgroups and interactions, and sensitivity analyses | Results; Fig. 4d; Supplementary Tables 2-4 and 7; Supplementary Figs 1-3 |
| Key results | 18 | Summarize key results with reference to study objectives | Discussion, paragraph 1 |
| Limitations | 19 | Discuss limitations, taking into account sources of potential bias or imprecision; discuss direction and magnitude of any potential bias | Discussion: limitations paragraph |
| Interpretation | 20 | Give a cautious overall interpretation considering objectives, limitations, multiplicity of analyses, results from similar studies and other relevant evidence | Discussion; Box 1 |
| Generalisability | 21 | Discuss the generalisability (external validity) of the study results | Discussion: limitations paragraph (self-selected, fee-paying users of a consumer platform) |
| Funding | 22 | Give the source of funding and the role of the funders for the present study and, if applicable, for the original study | Funding and Competing interests statements (end matter) |
